# Impact of Governmental interventions on epidemic progression and workplace activity during the COVID-19 outbreak

**DOI:** 10.1101/2020.06.05.20122903

**Authors:** Sumit Kumar Ram, Didier Sornette

**Affiliations:** ETH Zurich, Department of Management Technology and Economics Zurich, Switzerland; Institute of Risk Analysis, Prediction and Management (Risks-X) Academy for Advanced Interdisciplinary Studies Southern University of Science and Technology (SUSTech), Shenzhen, 518055, China

## Abstract

In the first quarter of 2020, the COVID-19 pandemic brought the world to a state of paralysis. During this period, humanity has seen by far the largest organized travel restrictions and unprecedented efforts and global coordination to contain the spread of the SARS-CoV-2 virus. Using large scale human mobility and fine grained epidemic incidence data, we develop a framework to understand and quantify the effectiveness of the interventions implemented by various countries to control epidemic growth. Our analysis reveals the importance of timing and implementation of strategic policy in controlling the epidemic. Through our analysis, we also unearth significant spatial diffusion of the epidemic before and during the lock-down measures in several countries, casting doubt on the effectiveness or on the implementation quality of the proposed Governmental policies.

## 1 Introduction

The pandemic due to the SARS-CoV-2 virus (*1*) has been impacting the world population, health care system and economies over the first half of 2020 (*2*). Since its identification in December 2019 in Wuhan, China, this novel coronavirus disease (COVID-19) has been spreading in China in Jan.-Feb. 2020. The epidemic development was detected in Italy in the second half of February and progressively diffused in the rest of the world. It was declared a global pandemic on March 11, 2020 by the World Health Organization (WHO) (*3*). We have all witnessed and experienced a series of interventions with various levels of confinement measures in different countries and regions, aimed at decreasing the effective reproduction number *R_t_* and controlling the epidemics.

Our starting hypothesis is that the development of infected cases, of the various forms of illnesses resulting from infection and of the death rates reflect the interplay between the biological and epidemiological properties of this new SARS-CoV-2 virus and the political, cultural, sociological and governance characteristics of different nations and human communities. By combining mobility data, epidemiological data and clinical data across ten countries, we develop a modelling framework to quantify the effectiveness of the interventions implemented by various countries to control the epidemic growth. This shock provides a real-life natural experiment to falsify the effectiveness of different organisations and interventions and policies, with the goal of informing and guiding future plans against potential second and third waves of the epidemics as well as future outbreaks in general.

First, we quantify precisely how interventions in the form of mobility restrictions and social distancing have had significant impacts in controlling the development of the epidemics in many countries, as measured by the decrease in reproduction number *R_t_* and the extend of curbing the increase of infected cases. We also document a surprisingly large heterogeneity in the reduction of *R_t_* across regions within a given country and also across countries. Further, we observe non monotonous and fluctuating time dependence of *R_t_* in different regions.

Our most surprising result is that, in many regions where the epidemic was not visible, the interventions led to a significant transient increase in reproduction number *R_t_*, in contradiction with the short-term objective of lockdown measures. For instance, in several regions of France and Italy, lockdown led to an obvious strong collapse of mobility, however accompanied by an increase of *R_t_*. We interpret this phenomenon as a result of preemptive large movements of people to relocate before the strict lockdown implementation, hence promoting new contagions and epicenters for the epidemics to mature for a while after the lockdown. Another plausible mechanism, which underlies the Japanese policy, is the effect of closed spaces and close-contact settings within confined households.

We also quantify how the timing of lockdown determined the trajectory of the epidemic by estimating the spatial correlation across regions within a country of the total increase of infected cases after the lockdown. In a number of countries, the epidemic started much before the lockdown date and was developing silently as revealed by the strong spatial diffusion of the epidemic after the lockdown. In other countries, the epidemic was better contained by intervention measures.

Our analysis overall suggests that the interventions may have not been optimal and that there are probably better alternatives to complete lockdowns. Our study reveals the importance of timing and targeting of interventions as a likely better strategy compared with undifferentiated lockdown. We observe belated intervention at the regional and local levels and hasty global lockdowns, which, in a number of regions, result in disappointing reduction of the reproduction number and curbing of the increase of new cases compared to other comparable countries.

## 2 Results

We use a probabilistic contagion model with inhomogeneous source terms to explain the temporal evolution of epidemic because of COVID-19 (*4*). Using the daily number of confirmed cases and the generation time model (see figure (S1)), we estimate the time evolution of effective reproduction number *R_t_* – which is defined as the actual average number of secondary cases per primary case – with a Sequential Bayesian estimation. As a typical result of our analysis, figure 1 shows the time evolution of the effective reproduction number in the *Hessen state* in *Germany*.

**Figure 1:**
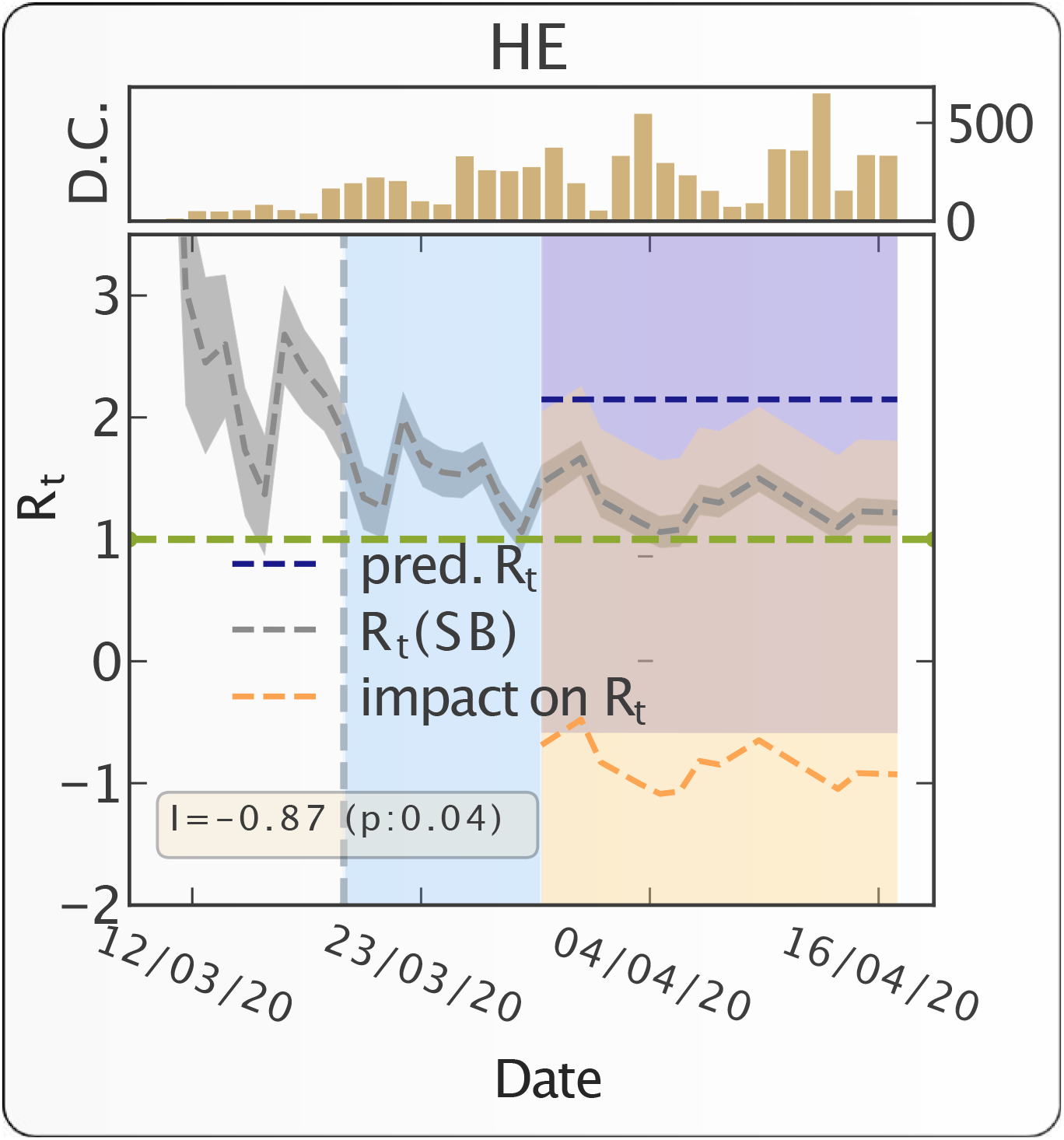
Progression of epidemic and impact of travel restrictions on epidemic growth in the Hessen state in Germany. The grey dashed line (with 95% CI band) represents the most likely estimation of *R_t_*. The dotted vertical line represents the start date of intervention. The region between intervention date and effective intervention date – 10 days following the intervention date – is marked with light blue color. It indicates the period during which people exposed to the virus prior to the intervention date would keep on appearing as the new confirmed cases. The counterfactual predicted *R_t_* is presented by the dashed blue line (with 95% CI band). The pointwise impact of the intervention is presented as the dashed orange line (with 95% CI band). The horizontal green line represents the critical value *R_t_* = 1. The inset figure shows the time evolution of number of daily confirmed cases (D.C.). We note down the absolute impact – average value of point wise impact – of the intervention along with the p-value in the yellow box. The p-value measures the probability of observing the impact by random chance.

With the help of a diffusion-regression state-space model (*5*) and MCMC posterior inference, we estimate the counterfactual evolution – that would have occurred had no intervention taken place – of post intervention effective reproduction number. The comparison between the effective reproduction number and the predicted *R_t_*, had no intervention taken place, allows us to quantify the effectiveness of intervention for the *Hessen state* in *Germany* shown in fig. 1. We find an average reduction of *R_t_* of 0.87, and reject the null hypothesis with a p-value of 0.04 that this reduction could result from chance under the counterfactual evolution without lockdown.

Figure 2 shows the map of the absolute impact on *R_t_* as a result of intervention for different states in Germany. The color for each state represents the average impact of the intervention on *R_t_*, i.e., the magnitude increment or decrement of *R_t_* from the counterfactual predicted value without lockdown over the thirty days following lockdown. The radial wedges represent the temporal evolution of *R_t_* in the corresponding state and color of the strips represent *R_t_* on a particular day over the thirty days following lockdown. This figure illustrates the significant heterogeneity in the results, as well as an important non-monotonicity in the dynamics of *R_t_*.

**Figure 2:**
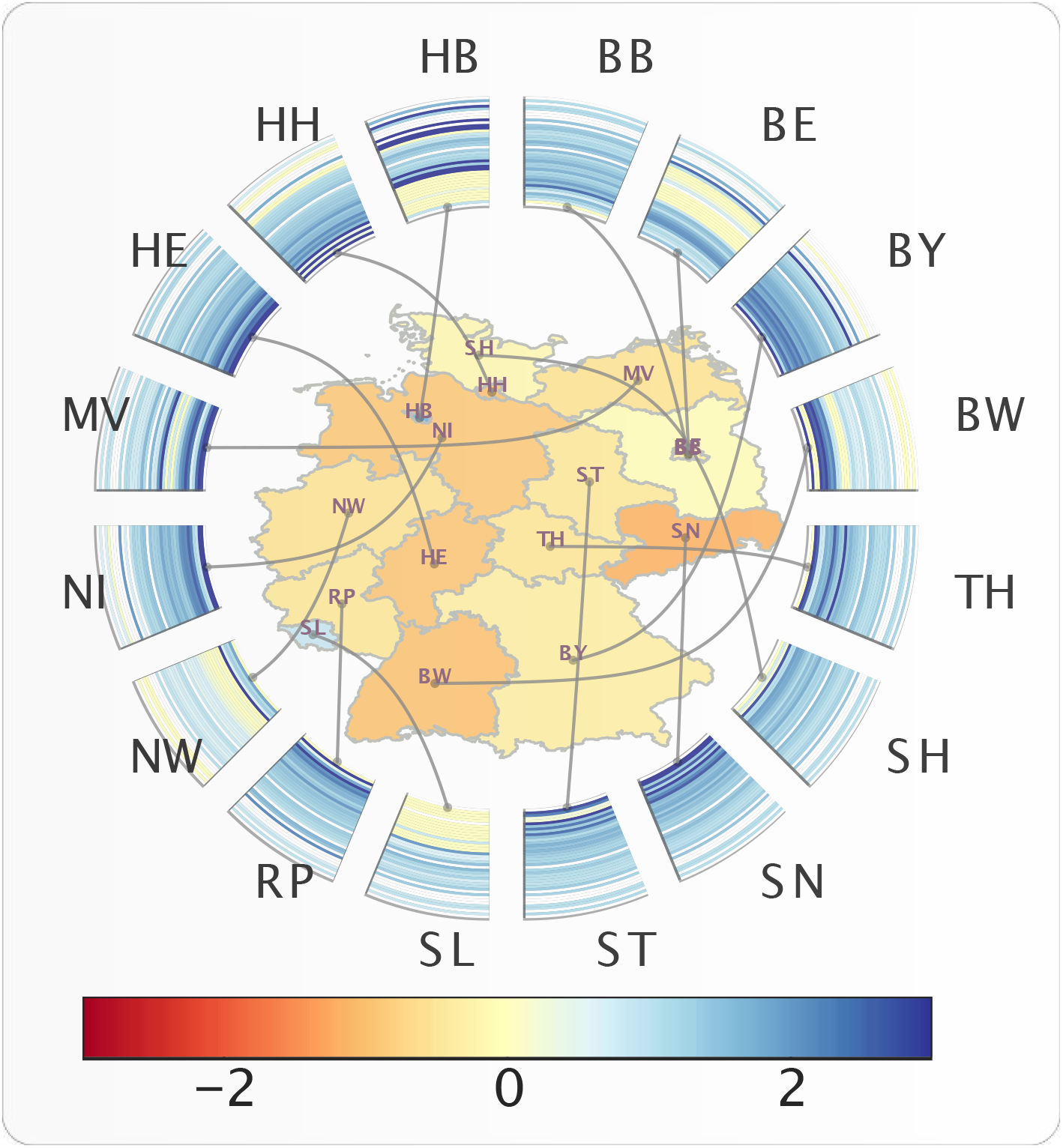
Time evolution of *R_t_* and absolute impact of travel restrictions on *R_t_* for different states in Germany. The color for each states (name in ISO 3166-2 code notation) represents the absolute impact (increase or decrease of *R_t_*) due to travel restriction in that state. The radial wedges represent the time evolution of *R_t_* in the corresponding state and color of the strips represent *R_t_* on a particular day.

We then analyse the impact of Governmental interventions on human mobility, illustrating the results for Germany. We are able to breakdown the impact of intervention in different mobility dimensions and quantify the level of reduction of mobility (see fig. 3 for details). Figure 4 shows the absolute impact on workplace activity (in % change from baseline) in different states in Germany, resulting from the intervention. The radial wedges represent the temporal evolution of the workplace activity in the corresponding state, and color of the strips represent activity on a particular day. Contrary to the map of the absolute impact on *R_t_* (figure 2), the absolute impact of travel restrictions on mobility is much more homogenous across German states, and also consistent along the time axis.

**Figure 3:**
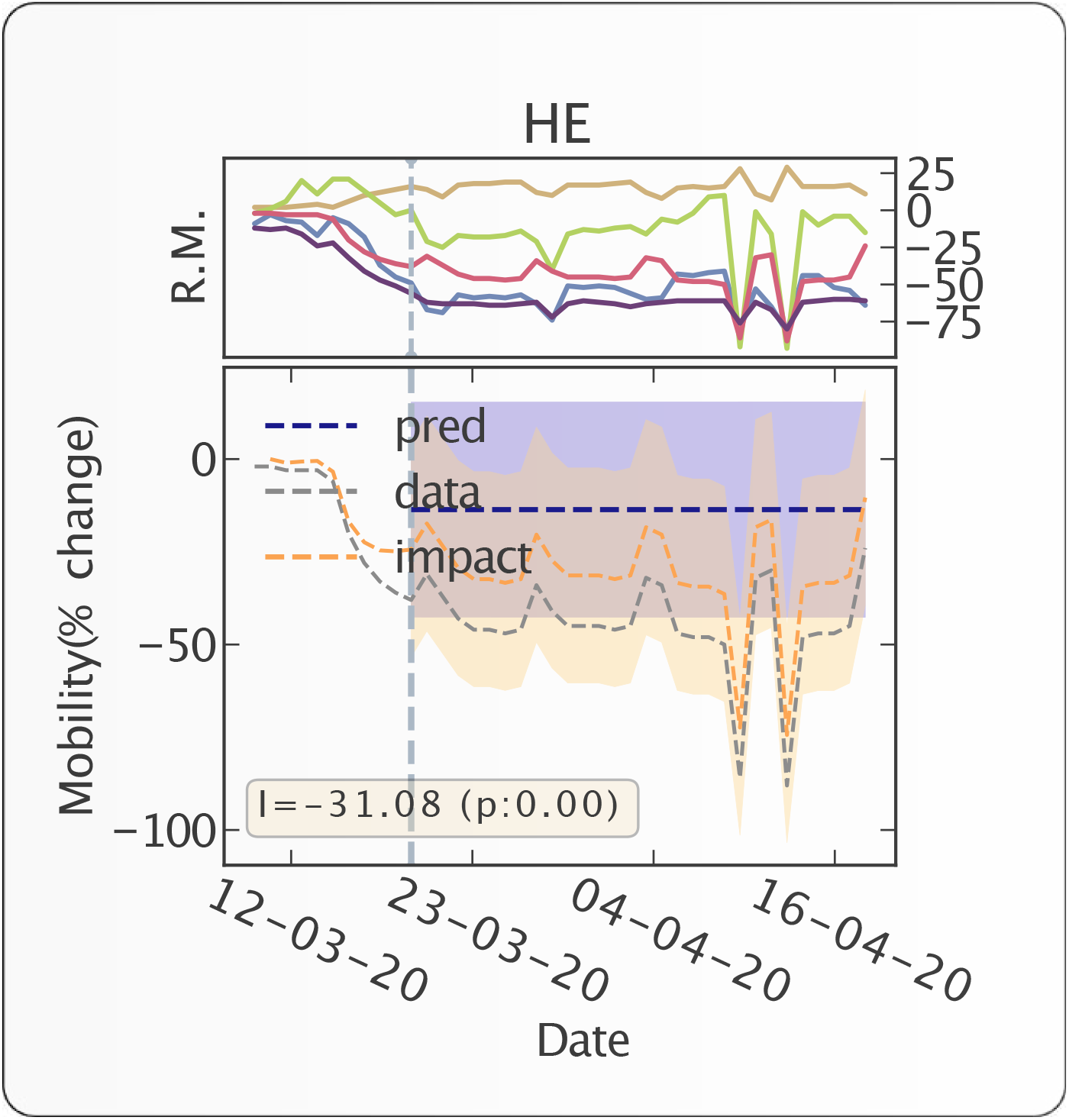
Countrywide time evolution of human mobility and impact of travel restrictions on human mobility for Germany. The top panel represents the time evolution of human mobility (% change from the baseline activity) in different mobility dimensions (yellow: home, red: work, blue: retail, green: grocery, violet: transit). Transit is a proxy for long-distance travel (it corresponds to petrol pumps/filling stations etc.). In the bottom panel, The y-axis represents % increase or % decrease of the average individual’s activity. the grey dashed line represents the % change of activity from the baseline (baseline is set to 0) in the workplace resulting from the intervention. The dotted vertical line represents the intervention date. The counterfactual predicted evolution of workplace activity, had no intervention taken place, is presented by the dashed blue horizontal line. The point-wise impact of the intervention on mobility is presented by the orange dashed line. The yellow box indicates the absolute impact of the intervention along with its p-value.

**Figure 4:**
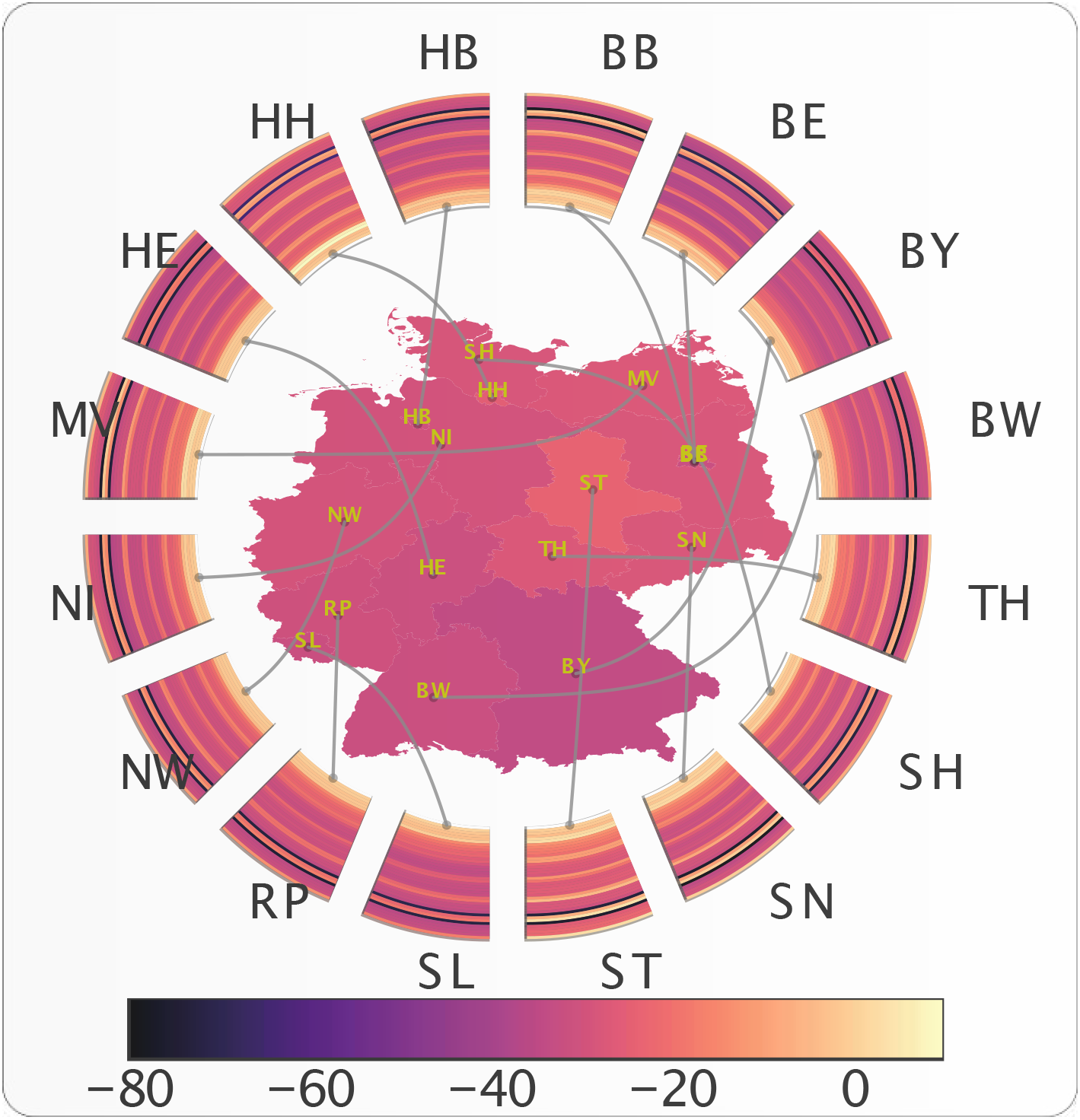
State-by-state time evolution of mobility and absolute impact of travel restrictions on mobility. The color of each state (name in ISO 3166-2 code notation) represents the absolute impact (increase or decrease of mobility) of interventions on workplace activity in that state. The radial wedges represent the time evolution of mobility in the corresponding state and color of the strips represent the mobility on a particular day. SL is Saarland, ST is Sachsen-Anhalt.

We then evaluate the impact of Governmental interventions on *R_t_* as well as on works place activity for the top level administrative divisions in a number of countries. Figure 5 illustrates the impact of Governmental interventions on workplace activity in a number of countries and figure 6 shows the impact on *R_t_*. Each subplot of figure 5 shows the time evolution of mobility and the impact of travel restrictions on mobility of different top level administrative divisions of a country. Each ring map represents a country and provides a visual representation of the time evolution of work place activity as well as the impact of Governmental interventions on the workplace activity. Each subplot of figure 6 shows the time evolution of the effective reproduction number *R_t_* and the absolute impact of travel restrictions on *R_t_* in ten countries. Each ring map represents a country and shows the time evolution of the effective reproduction number *R_t_* as well as the impact of Governmental interventions on *R_t_*. Figure 5 shows a rather homogeneous impact on workplace activities across the different regions of each of the eight analysed countries. However, there is a large heterogeneity across different countries, e.g. Spain being the most effected country while Japan is the least effected country in terms of workplace activity. In contrast, the impact on *R_t_* is quite heterogeneous across different regions within a country, notwithstanding similar levels of restriction, as illustrated in figure 6 by Kerala (KL) and Maharastra (MH) in India. Surprisingly, across a number of regions, a significant increment of *R_t_* is observed following the implementation of very strict lockdowns (e.g. Maharastra (MH) in India and Luzern (LU) in Switzerland.).

**Figure 5:**
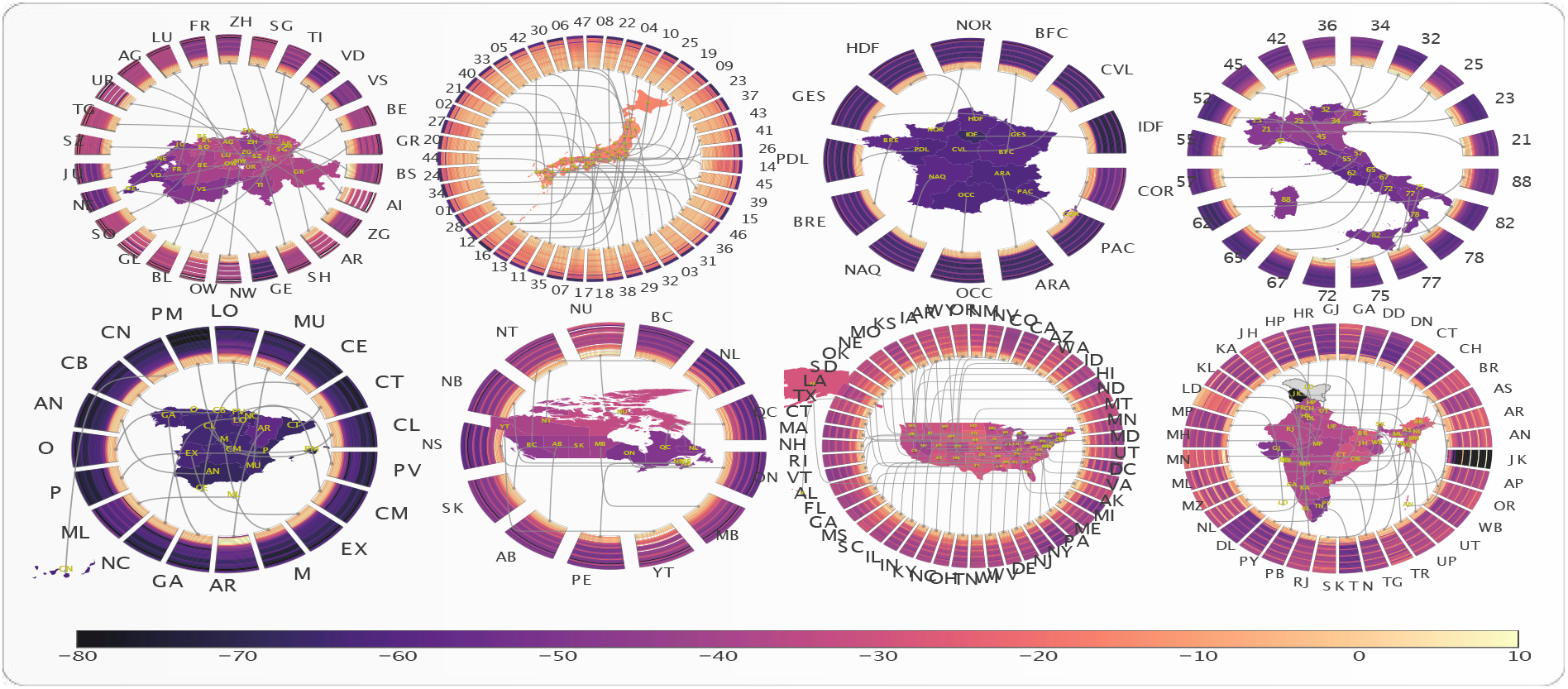
Time evolution of mobility and impact of travel restriction on mobility in eight countries. Each subplot shows the time evolution of mobility and impact of travel restrictions on mobility of different top level administrative divisions of a country. The countries are, for top left to bottom right: Switzerland, Japan, France, Italy, Spain, Canada, USA and India. The color of the regions on the map denoted by their ISO 3166-2 code represent the impact (increase or decrease of mobility) of travel restriction on that region. The radial connected wedges represent the time evolution of the mobility in the corresponding region or state for each country. The color of the strips in the wedges represent the mobility on a particular day.

**Figure 6:**
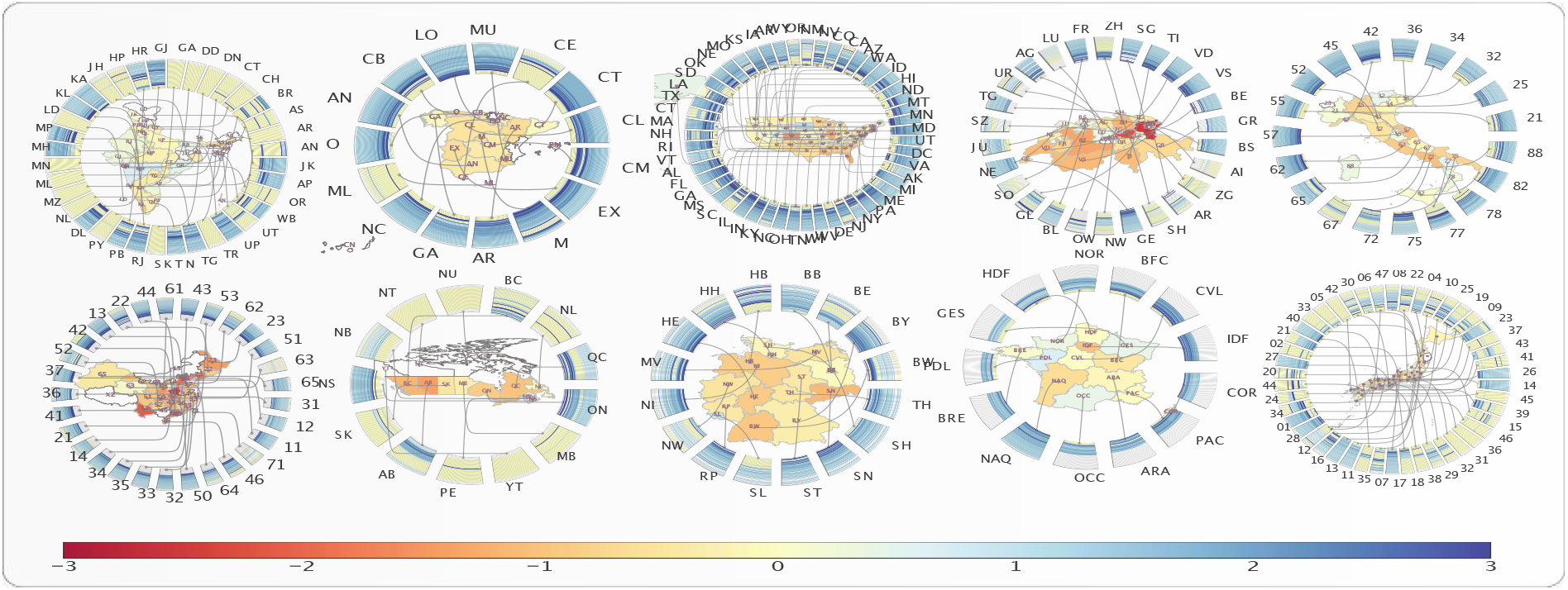
Time evolution of effective reproduction number (R_t_) and absolute impact of travel restrictions on *R_t_* in ten countries. Each subplot presents time evolution of *R_t_* and impact of interventions on *R_t_* for different top level administrative divisions of the country. The color of the regions (name in ISO 3166-2 code notation) represents the impact (increase or decrease of *R_t_*) of interventions on that state. The radial wedges represent the time evolution of *R_t_* in the corresponding state, and color of the strips represent *R_t_* on a particular day. The countries are, from top left to bottom right: India, Spain, USA, Switzerland, Italy, China, Canada, Germany. France and Japan.

Figure 7 shows the joint distribution (obtained by kernel density estimation) of the absolute impact resulting from intervention on workplace activity and on the *R_t_* in the administrative divisions of nine different countries. In other words, figure 7 compares the strictness of the governmental interventions, measured in terms of the impact on the workplace activity, against the corresponding reduction/increment in effective reproduction number. There is no significant correlation between these two variables, suggesting that other variables are controlling the reduction of reproduction rates.

**Figure 7:**
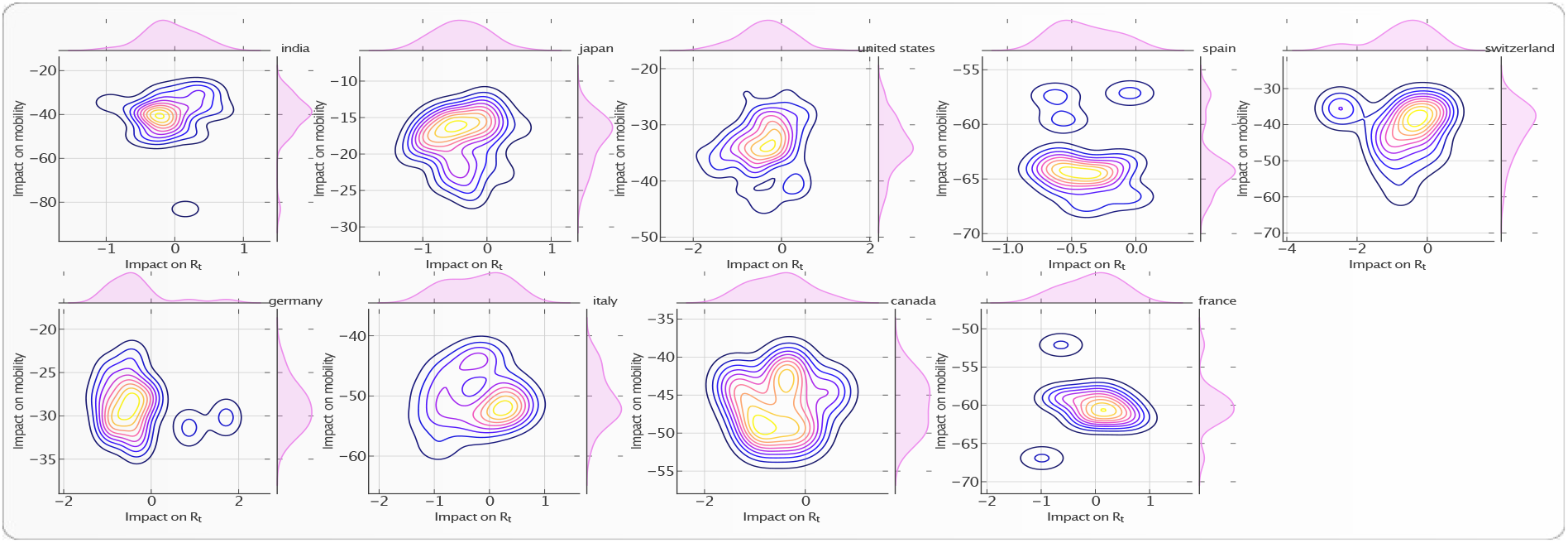
Impact of travel restrictions on workplace activity and on epidemic progression (R_t_) in nine countries. Each panel represents the bivariate kernel density estimation as a function of the absolute impact on workplace activity and impact on *R_t_* in the administrative divisions of each country. The bivariate distribution is constructed over the set of regions within each country. The top and right inset of each of the nine plots represent the marginal distribution of the respective variables for each country. The countries are, from top left to bottom right: India, Japan, USA, Spain, Switzerland, Germany, Italy, Canada and France.

Figure 8 represents the spatial correlation analysis of the total increase ∆*S* in the number of confirmed cases during the first 30 days of interventions. We define *Moran’s I*, which is a measure of spatial auto-correlation, as follows:

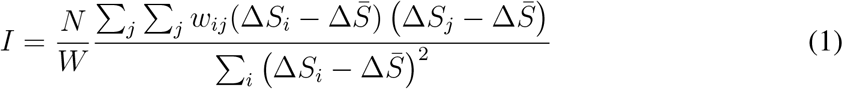

where *N* is the number of spatial units indexed by *i* and *j* in a given country; Δ*S* is the increase in the number of confirmed cases; 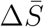 is the mean of Δ*S* over all the spatial units of the country; *w_ij_* is a matrix of spatial weights with zeroes on the diagonal (i.e., *w_ii_* = 0); and *W* is the sum of all *w_ij_*. We define *W* by giving a weight *1* if two regions are neighbors, and *0* otherwise. The “spatial lag” of Δ*S* for a region is defined as the weighted sum of its neighbors’ Δ*S*. The scatter plot in each panel of figure 8 shows the spatial lag as a function of its corresponding Delta S for different regions in each country. The inset gives the kernel density estimation of the simulated Moran’s I from the null model of no spatial correlation. The slope of the scatter plot of Δ*S* against the spatial lag is known to converge to the *Moran’s I* (*6*). We test the significance of the *Moran’s I* under the null hypothesis of no spatial auto-correlation and simulate *1000* realizations, by randomly shuffling the locations of the Δ*S*. The spatial correlation analysis presented in figure 8 reveals significant spatial correlation of Δ*S* in Italy, Switzerland, Japan and United States.

**Figure 8:**
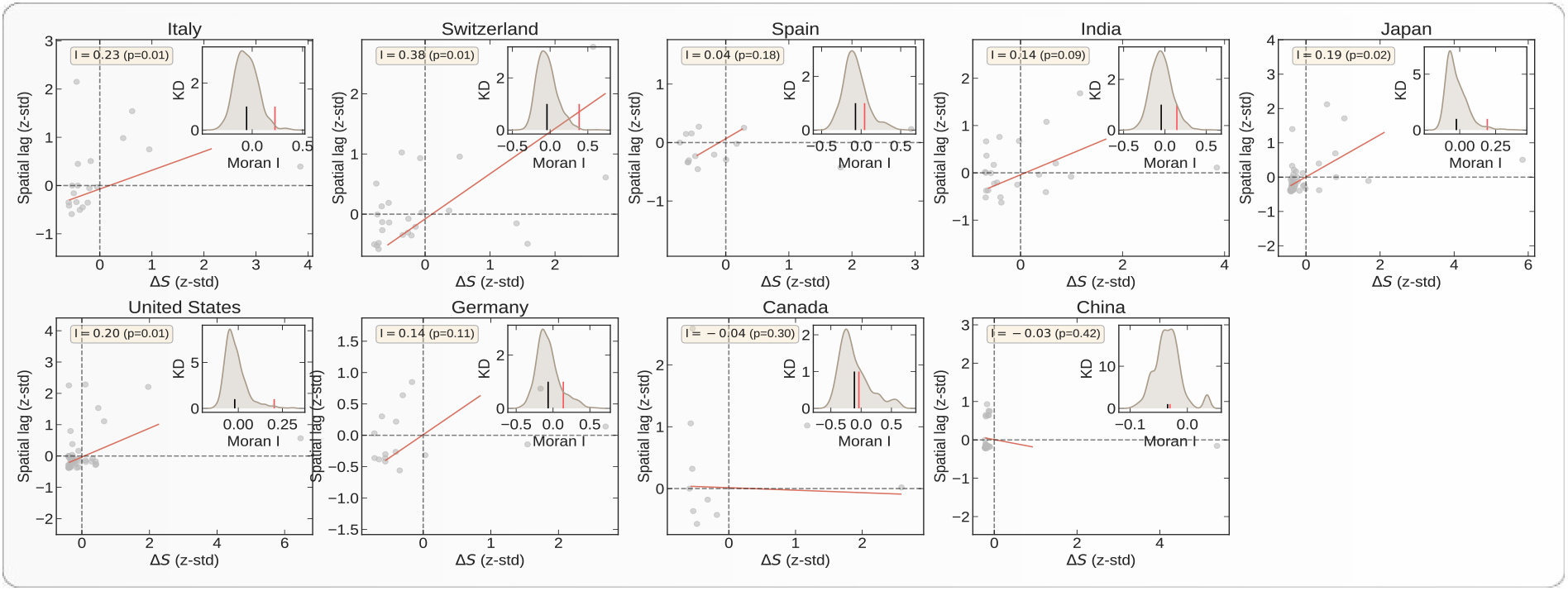
Spatial auto-correlation of the total increase Δ*S* in number of confirmed cases during the first 30 days of intervention in nine countries. In each panel, the x-axis corresponds to the value of Δ*S* in a given region in a given country; the y-axis gives the average Δ*S* over the neighboring regions, called “spatial lag” in the caption along the y-axis. These two variables are *z-standardised* for better comparison. The inset in each panel represents the Kernel Density estimator for the distribution of the simulated *Moran’s I*. The black vertical line in the inset represents the expected *Moran’s I* from simulations with the null hypothesis of no spatial correlations. The red vertical line represents the value obtained from empirical data. *Moran’s I* along with its p-value is given in yellow box. The countries are, from top left to bottom right: Italy, Switzerland, Spain, India, Japan, USA, Germany, Canada and China.

In order to understand the effectiveness of lockdowns, we compare the number of confirmed cases against the impact on *R_t_* during the first 30 days of interventions. Figure 9 shows the kernel estimation of the bivariate distribution of the total increase in number of cases and of the total impact on *R_t_* during the first 30 days of intervention in different regions of nine countries. The inset in each panel represents the impact on *R_t_* during this period, (or Δ*R_t_*), against the average *R_t_* before the intervention (or 〈*R_t_*〉*_init_*) over the regions in each country. The kernel density estimation of the bivariate distribution in figure 9 reveals a negative correlation between the above variables, showing a slowdown of the epidemic growth notwithstanding the increase of *R_t_*. We also compare the average *R_t_* before the intervention (or 〈*R_t_*〉*_init_*) and impact on *R_t_* (or Δ*R_t_*) from the intervention to understand the effectiveness of intervention in reducing/increasing *R_t_* from its initial values. The inset panels of figure 9 unveil a negative correlation between these two variables, indicating a significantly large decrease of *R_t_* in the regions of larger initial *R_t_*. Surprisingly, we also see the extension of the distribution to the second quadrant, revealing the fact that, in many places, the epidemic started after the lockdown.

**Figure 9:**
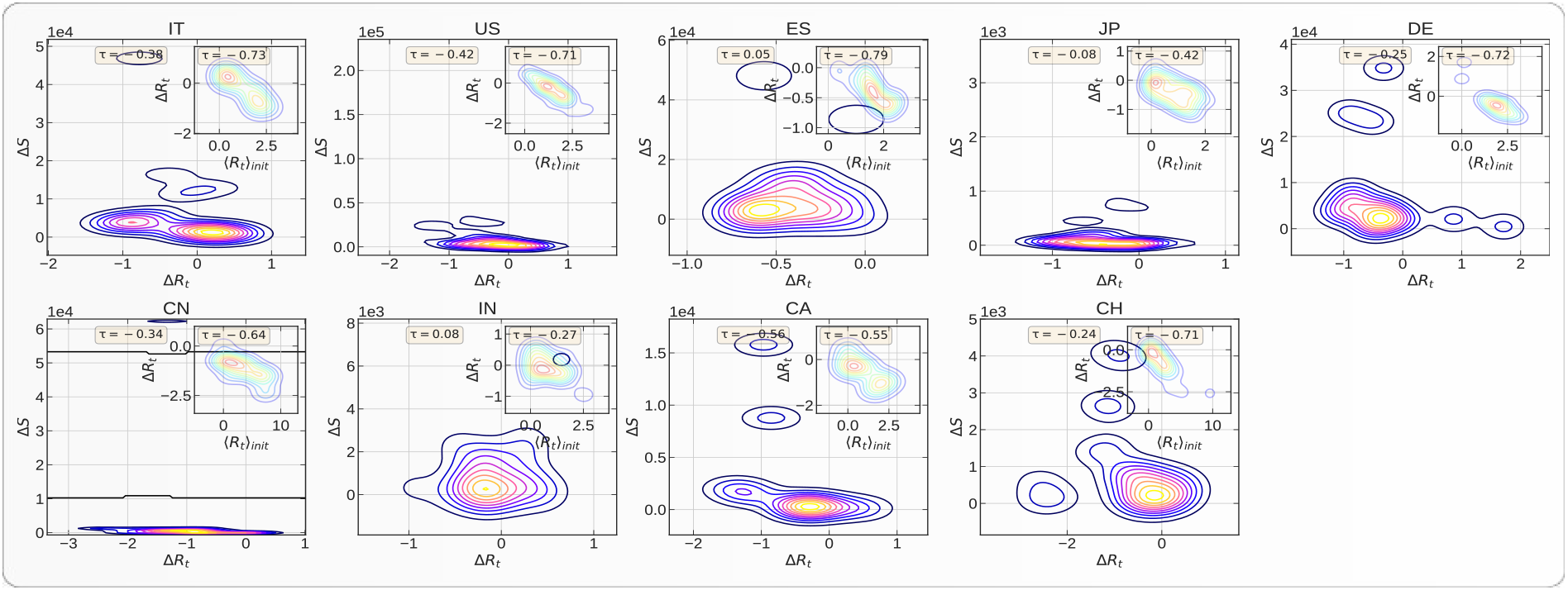
Joint distribution of the total number Δ*S* of confirmed cases within the first 30 days of intervention and of the absolute impact of intervention on the effective reproduction number (Δ*R_t_*) within this period. Each panel represents the Kernel Density Estimation for the total number of confirmed cases within the first *30* days of the intervention against the impact of interventions on effective reproduction number (Δ*R_t_*) in different administrative divisions of a country. The bivariate distribution is constructed over the set of regions within each country. The yellow box contains the Kendall *τ* correlation value for this joint distribution. The inset in each panel represents the variation of Δ*R_t_* during the intervention against the average initial *R_t_* before the intervention. The yellow box gives the corresponding Kendalls *τ*’s. The countries are, from top left to bottom right: Italy, USA, Spain, Japan, Germany, China, India, Canada and Switzerland.

**Figure 10:**
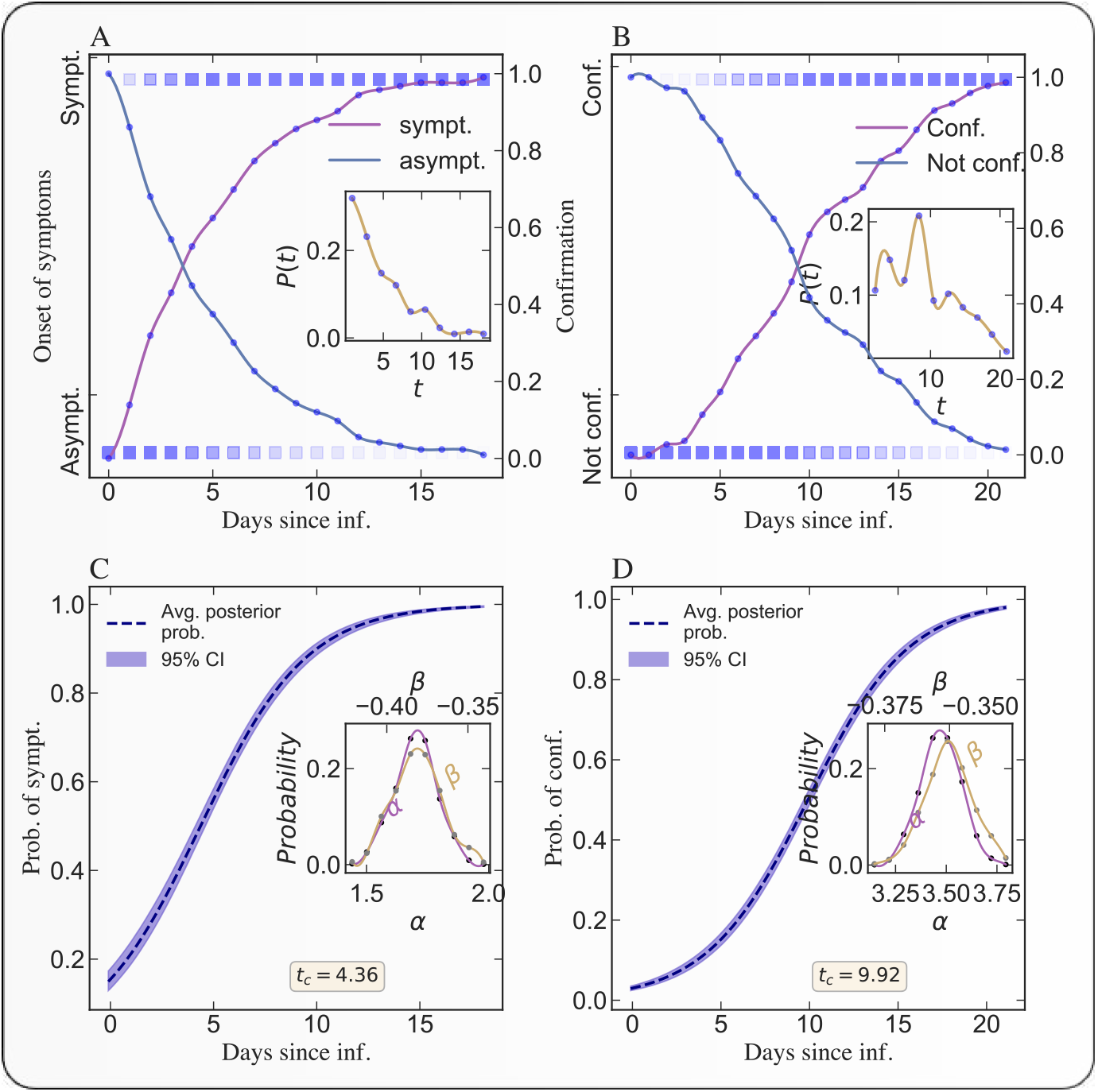
Bayesian inference of the incubation period and confirmation period. A) Empirical cumulative probability of developing a symptom on a particular day following the day of exposure to the virus. We use individual level clinical data (*8*, *9*) to conduct these Bayesian estimations. The lighter color of the square represents low probability and the deep color represents high probability. The solid lines represent the fraction of people remaining asymptomatic (decreasing curve) or becoming symptomatic (increasing curve) on a given day. following exposure. The inset figure represents the empirical probability distribution of duration between the date of exposure and the date of onset of symptom or the clinical confirmation. B) Same as A) but for the probability of being confirmed. C) Estimation of the posterior probability distribution (with 95% confidence interval) of the incubation period (i.e., of getting the clinical confirmation on *day* = *t*, provided the individual is exposed to the virus on *day* = 0). The dashed solid line represents the most likely posterior probability estimation and the light band represent the 95% confidence interval. The inset figure presents the distributions of the two estimated parameters in the logistic model (*2*). D) Same as C) for the probability distribution of confirmation period. The median value (and also mode) of the incubation period is 4.38 days. The median value (and also mode) *t_c_* of the confirmation period is 9.94 days.

The tables S1 to S10 in the supplementary materials provide the detailed values of *R_t_* with positive and negative impact along with the corresponding growth in total number of confirmed cases as a result of intervention in the different regions for each country. The regions that are characterised by an increase of *R_t_* after lockdown are indicated in bold face.

### 3 Discussion

We have found important to quantify the incubation and confirmation periods in order to assess the impact of policy interventions to contain the epidemics in different regions. This quantification shown in figures 10 and S1 allows us to define a credible time interval over which to quantify the impact of interventions. With this, our analysis provides a framework to under- stand the effectiveness of the policies and interventions implemented by various countries to control the epidemic growth. The wealth of results presented in figures 1–9 leads to the following insights.

Overall, it is clear, and unsurprising, that interventions in the form of mobility restrictions and social distancing are found to have significant impacts in controlling the development of the epidemics in many countries. In particular, we have quantified the decrease in reproduction number *R_t_* resulting from the intervention. As an illustration shown in fig. 1, for the Hessen state in Germany, we are able to quantify that intervention in this state reduced *R_t_* by about 1 unit compared with the counterfactual scenario of no intervention.

More surprising is the large heterogeneity in the reduction of *R_t_* across different states in Germany, as shown in figure 2. For this country, intervention measures had led systematically to a decrease in *R_t_*, with quite strong differences from state to state. Even more surprising is the time dependence which exhibits a non monotonous and fluctuating behaviour of *R_t_* in different regions.

For most of regions in different countries, because of the interventions, *R_t_* decreased, however there are some places where it increased. Because this increase is transient and constrained by the lockdown, it does not lead to a very strong explosion of new cases. A tentative interpretation is that, as lockdown was considered and being implemented, in a number of regions, it triggered large movements of people to relocate, hence promoting new contagions and epicenters for the epidemics to mature after the lockdown. An additional mechanism, which underlies the Japanese policy, is the effect of closed spaces and close-contact settings within confined households, which has been shown to lead to increased infections within households for instance. But because the lockdown only allows the new nucleii of contagion to develop locally, the number of cases did not explode. Examples of this effect can be found in Luzern and Solothurn of Switzerland, in Bremen of Germany, in Saga Ken of Japan and in Odisha of India. This effect that has previously been described qualitatively is given here a quantitative support by our systematic analysis.

The observation that lockdown led first to an increase in *R_t_* in a significant number of regions and countries is confirmed by the correlation analysis presented in fig. 9 relating increase of cases to changes of *R_t_* in different countries. There are many regions in each country where there was no epidemics before the lockdown. The lockdown triggered a pre-emptive movement of people to relocate, increasing *R_t_* after the lockdown. But the increase of *R_t_* did not increase Δ*S* too much due to the effect of confinement. The numerous local infectious cases could only infect their immediate relatives. In regions where the epidemic was at more advanced stages, the lockdown had the effect of decreasing *R_t_*, as expected. The insets in fig. 9 provide further support for this conclusion. The average correlations shown by the plots indicate that, where there was no epidemic, the epidemic started after the lockdown, and where there was an on-going epidemic, it came under control.

We also quantify that there is not much advantage resulting from strict lockdowns. For example, Japan and Switzerland did rather well in spite of weaker lockdowns, whereas Italy and Spain performed much more poorly with stricter confinements. This is related to the effectiveness of early stage contact tracing as well as effective awareness of the role of protection measures. Figure 7 exemplifies this point by showing the paradoxical results that lockdown led to an obvious strong collapse of mobility accompanied by an increase of *R_t_* in several regions in France or Italy, for instance. For the other countries, the effect of lockdown is more as expected.

Moreover, the timing of lockdown is very important in determining the trajectory of the epidemic. Figure 8 shows the spatial correlation of the total growth of epidemic after the lockdown. The larger is the spatial diffusion (possible during the advanced stage of the epidemic), the larger is the spatial correlation. Our analysis shows that, for Italy, Switzerland, the United States, and Japan, the spatial correlation of Δ*S* is significantly positive. This means that, in these countries, the epidemic started much before the lockdown date and was developing silently as revealed by the strong spatial diffusion of the epidemic. While Switzerland and Japan contained the epidemic with effective containment policy, Italy and United States failed to do so because the intervention was ill-adapted to the spatial developments.

When comparing the German, Swiss and US lockdowns via their mobility data, we find very similar severity levels of the confinements. However, the effectiveness of the lockdown to control the epidemic in the USA is quite low, while very significant in Germany (*7*) and Switzerland. While Switzerland and USA both imposed the lockdown at a rather late stage of their unfolding epidemic, the Swiss containment and awareness policy was significantly superior to that in the USA. We quantify that the epidemics has diffused to many states in the USA, as revealed by the spatial correlation in figure 8, even after the lockdown was implemented. This explains the failure to reduce the transmission to a large degree. For Italy and Spain, because there was a significant numbers of confirmed cases across different regions of Italy and Spain before the lockdown, it is difficult to determine the effectiveness of cross-region transmission control.

In most of the Indian states, we do not observe any significant impact of the lockdown on reducing the effective reproduction number. A possible explanation is that, in most of the places, there is no real epidemic and the majority of the infection cases are found in a small number of states. The complete lockdown of the entire country might have been ignorant to this very strong heterogeneity.

We also unearth some outliers, e.g, Maharastra state in India, Saarland in Germany where, despite a strict lockdown (quantified in our analysis by a strong reduction of workplace activity), both the number of cases and *R_t_* exploded. This poses the questions of unobserved contagion paths, likely associated with specific events, perhaps the existence of super-spreaders and so on.

Our analysis overall suggests that the interventions may have not been optimal in many countries and that there are probably better alternatives to complete lockdowns. One alternative is a sequential and selective lockdown approach, putting in selective quarantine based on a threshold value for the number of confirmed cases while leaving the other places more open with social distancing but not complete lockdown. One should however stress that this alternative intervention requires very strong testing support in order to determine with sufficient reliability the positive cases. The case of India supports further the idea that the policy was too early in implementing a complete lockdown of the country for such a long time but too late implementing effective quarantine of people coming from effected places. The cases of Italy, France and other regions where confinements led to a transient increase of *R_t_* over the following 30 days also underlies the plausible importance of controlling close contacts and confined places: with the aim of doing good, confining might have worsen the transmission of the disease in a number of cases.

### 4 Materials and Methods

#### 4.1 Data

##### 4.1.1 Clinical Data

###### Incubation period and confirmation period

To calculate various epidemiological parameters, we use the individual-level data (*8*, *9*) on COVID-19 epidemic which are geo-coded and include symptoms, key dates (date of onset, admission, and confirmation), and travel history. The dataset is curated from different sources, including official government sources (official websites of Ministries of Health or Provincial Public Health Commissions), peer-reviewed scientific articles, online reports, and news websites. From this database, we select 216 individuals, who got a clinical confirmation for covid-19 and were symptomatic as well as traveled to suspected places of exposure. We have the exact dates of travel, onset of symptoms, and of clinical confirmation for these 216 individuals.

###### Serial interval

We use the infector-infectee pairs dataset that has been collected from publicly available information published in research articles and quoted from official reports of outbreak investigations (*10*). We use 28 probable pairs for the estimation of generation time. For these 28 pairs of individuals, the date of illness onset for the pairs of individuals are defined as the date on which a symptom relevant to COVID-19 infection appeared and are determined by the reporting governmental body.

##### 4.1.2 Epidemiological Data

We collect the daily reported cases at the first administrative level divisions for a list of countries from various sources. The primary sources of information for the datasets are often local news agencies, government reports, WHO reports, various medical communities. Table 1 gives a list of countries with their starting date of the Governmental interventions. There is no lockdown in Japan like other countries, however, prime minister Abe on 7 April, proclaimed a state of emergency. This was the first emergency declaration in Japan and we consider this date as the starting date of intervention.

**Table 1:**
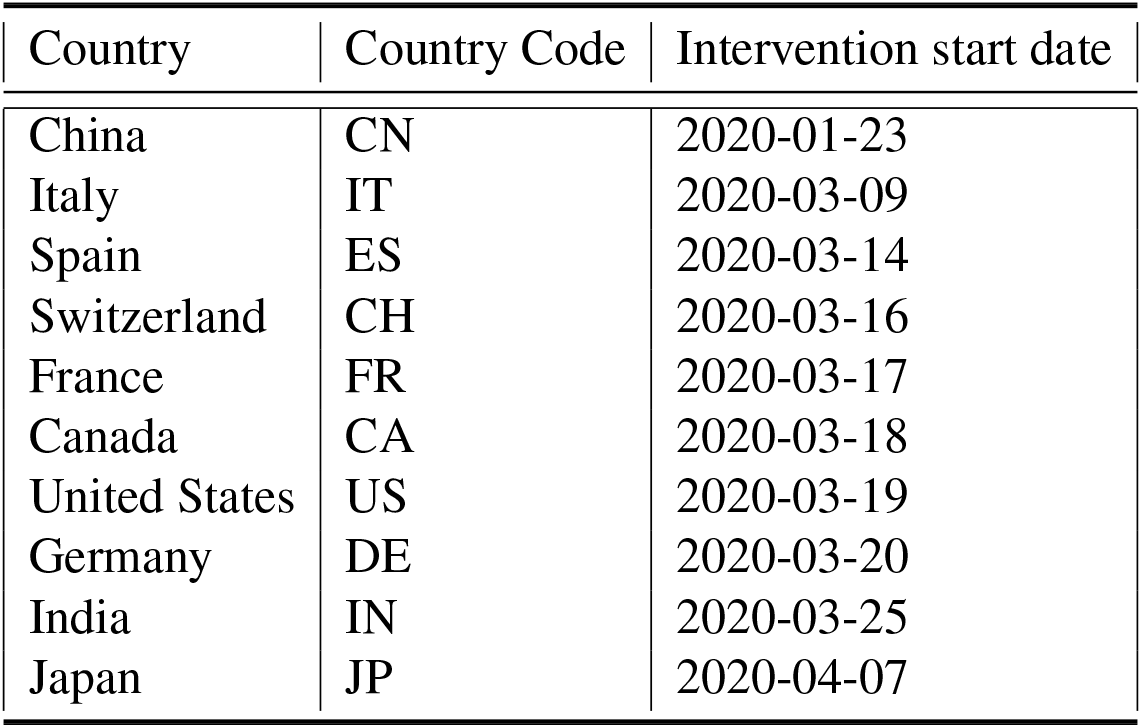
Starting dates of Governmental interventions (includes the lockdown) for controlling the outbreak. (source: https://en.wikipedia.org/wiki/COVID-19_pandemic_lockdowns)

##### 4.1.3 Mobility Data

We use the aggregated anonymised community mobility dataset that has been collected through the Google Maps app (*11*), to help understand what has changed in response to the Govt. policies aimed at flattening the curve of the COVID-19 pandemic. The dataset is anononymised to ensure that no personal data, including an individual’s location, movement, or contacts, can be derived from the resulting metrics. The anonymization process for the data includes differential privacy (*12*), with intentionally added random noise to metrics in a way that maintains both users’ privacy and the overall accuracy of the aggregated data (*13*). The dataset contains the percentage changes of the anonymized mobility metrics of Google users from a baseline based on the historical part.

##### 4.1.4 Map Data

We collect the geojson files for the first-level administrative divisions of the list of countries containing the names of the regions and the geometry from a number of openly accessible github repositories.

#### 4.2 Models and methodology

For our analysis, we select a time span that starts 10 days before the implementation of Governmental interventions – for controlling the outbreak, often through mass lockdowns – and ends 30 days following the starting date of Governmental interventions.

#### 4.2.1 Epidemiological estimations

The time between exposure and onset of symptoms is defined as the incubation period. We use a Bayesian framework to model the day of onset of symptoms following the date of exposure to the virus and the day of clinical confirmation of the virus following the date of exposure. Using the key dates for the selected 216 individuals (*8*, *9*), we estimate the important epidemiological parameters like incubation period, confirmation period (the number of days it take to get the clinical confirmation about the virus following the date of exposure). We use the logistic model

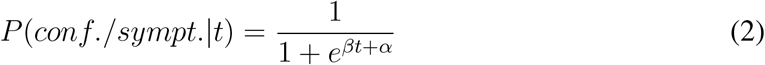

to estimate the cumulative probability distribution of individuals developing symptoms or getting the clinical confirmation on the *day* = *t* that they are infected, conditional on the fact that the individual was exposed to the virus on *day* = 0 and will be eventually symptomatic and will get a clinical +*ve* confirmation result. With the help of Metropolis-Hastings algorithm (*14*), – a Markov Chain Monte Carlo (MCMC) method (*15*)– we sample the model parameters (α, β), from a standard normal prior and train our model on the dataset to estimate the posterior probability distribution of developing symptoms or getting the clinical confirmation on the *day* = *t*, following the day of exposure on *day* = 0.

#### 4.2.2 Estimation of the time-dependent effective reproduction number *R_t_*

The principal epidemiological variable characterizing a disease’s transmission potential is the basic reproduction number, *R*_0_, which is characterized as the estimated number of secondary cases caused by a typical primary case, in an entirely susceptible population. When infection is spreading across a population, working with the effectively reproductive number *R_t_*, which is defined as the actual average number of secondary cases per primary case, is often more convenient. *R_t_* is normally smaller than *R*_0_, which reflects the impact of epidemic controls and the decline of susceptible individuals. The value of *R_t_* is comparable to the branching ratio in the Hawkes process (*16*, *17*), where *R_t_* > 1 can lead to an explosive growth of the epidemic and an ever-increasing number of new cases, while *R_t_* < 1 leads to the eventual demise of the growth process. Provided the branching structure describing who infected whom is determined, it becomes trivial to estimate *R_t_*. However, this information is not generally available and the estimation of *R_t_* then becomes tricky. Nevertheless, recent advances in epidemiology has made it possible to estimate the time evolution of the effective reproduction number (R_t_) through the observed epidemic curve in a geographical region (*4*, *18*). We use the method introduced by (*4*), which uses the daily number of confirmed cases and a model described below for the generation time (which is one of the key parameters dictating the severity of epidemic growth) to estimate the temporal evolution of effective reproduction number (R_t_) with the help of a Sequential Bayesian estimation approach.

The generation time is defined for source-recipient transmission pairs as the time between the infection of the source and the infection of the recipient. Because time of infection is generally not known, the generation time is often approximated by the serial interval, which is defined as the time between the onset of symptoms of the source and the onset of symptoms of the recipient. For the present case, we use the data for the serial intervals from (*10*), which has been constructed using the publicly available data. We calibrate the data against three models, i.e., Weibull, Log-normal, Gamma distributions to find the best possible approximation model from which the generation time is determined.

We use a probabilistic contagion model with in-homogeneous source terms to explain the progression of the COVID-19 epidemic (*4*). We consider both the human to human transmission and infections from the reservoir (contaminated surfaces) to explain the epidemic growth. Denoting *S*(t) and *N*(t) as the average number of susceptible and total population at time *t* and *β* and *γ*^−1^ as the contact rate and the infectious period, then

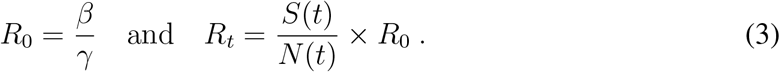

According to this model, if we denote the number of new infections from the reservoir between *t* and *t* + *τ* by Δ*B*(*t*) and the number of new cases within this period by Δ*T* (*t* + *τ*), then the stochastic discrete variable Δ*T*(*t* + *τ*) is generated by a probability distribution with average number of cases given by

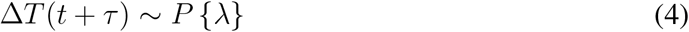

with

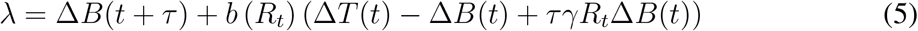

where *P* {*λ*} denotes a discrete probability distribution with mean *λ*. In eq. (*5*), *b*(*R_t_*) can be expressed as

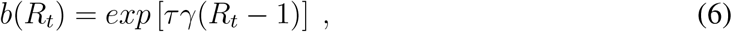

which is the slope of the tangent at the origin of case trajectories from the epidemic time delay plot (Δ(*t*) vs Δ(*t* − *τ*)) of surveillance data. More details about the derivation of the model can be found supplementary material or in (*4*).

We use a Bayesian framework to estimate the full probability distribution for the effective reproduction number *R_t_*, conditional on the time series for new cases. The probability distribution of *R_t_*, compatible with the observed temporal data stream, is given by

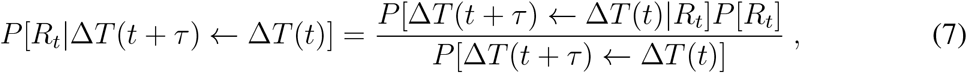

where *P* [*R_t_*] is the prior distribution and *P* [Δ*T* (*t* + *τ*) ← Δ*T*(*t*)] is independent of *R_t_*, and is a normalization parameter. From successive applications of Bayes’ theorem, a sequential estimation scheme, that uses streaming epidemiological observations performed in real time, can be constructed using the posterior distribution for *R_t_*, at time *t* as the prior in the next estimation step at time *t* + *τ*, leading to an update scheme via iteration of eq. (7). The resulting probability distribution for *R_t_* includes information on all observations up to time *t*, and thus is a robust estimator of the effective reproduction number compared to the estimation by only considering the cases between *t* and *t* + *τ*. Any changes in *R_t_* over time result from the assimilation of each new data point, leading to an updated estimate of *R_t_*. This in turn allows us the use of the estimation procedure as an anomaly detection tool.

#### 4.2.3 Estimation of the causal impact of Governmental intervention

We use the estimated time evolving effective reproduction number and the time evolving mobility metric to study the causal impact of various Governmental interventions to contain the spread of COVID-19. We use the framework proposed by (*5*) to infer the causal impact on the epidemic progression as well as the human mobility because of various Governmental interventions. The method uses a diffusion-regression state-space model that predicts the counterfactual evolution of effective reproduction number *R_t_*, as well as the mobility metric in a synthetic control that would have occurred, had no intervention taken place. The model is successful in inferring the temporal evolution of attributable impact, and flexibly accommodates multiple sources of variation, including local trends, seasonality and the time-varying influence of contemporaneous covariates by incorporating empirical priors on the parameters in a fully Bayesian framework. Using the MCMC for posterior inference, we estimate the most likely counter-factual evolution of effective reproduction number and the mobility metric during the first 30 days of the interventions.

According to the model, the generalized Bayesian Structural Time Series – state-space models for time-series data – can be expressed by

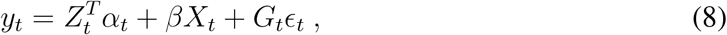

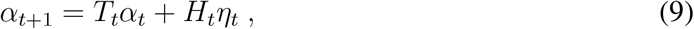

where 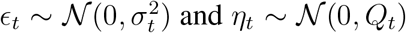 are independent of all other unknowns, *α_t_* is referred to as a “state” of the series and *y_t_* is a linear combination of the states plus a linear regression with the *covariates X*. Eq. 9 is the state equation governing the evolution of the state vector *α_t_* through time, whereas *y_t_* is a scalar observation. It is possible to model several distinct behaviors for the time series (including *ARMA* or *ARIMA*) by varying the matrices 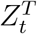, *T_t_*, *G_t_*, *Q_t_* and *H_t_*.

Here, we simplify (9) by taking Tt = Ht = 1, so that

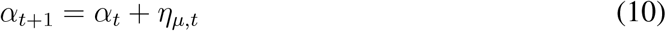

is a simple random walk, also referred to as the “local level” component. This random walk component embodies the increasing uncertainty of observations as time passes. In (*8*), we take 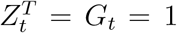 but augment the equation by accounting for the possible presence of seasonal components embodied into *γ_t_* described below. This allows us to reduce expression (*8*) into

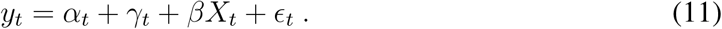

The seasonal components in eq. (11)) can be expressed as

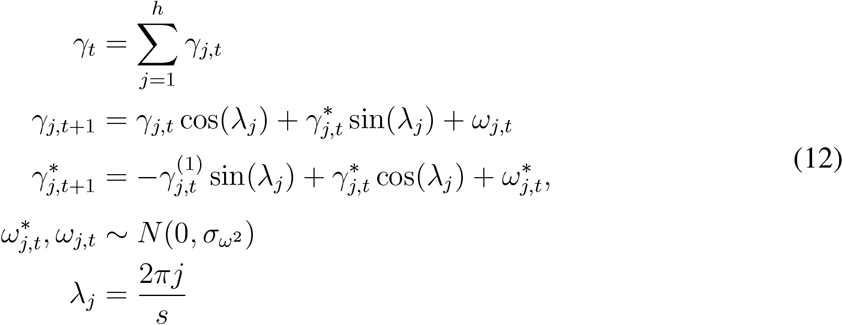

The linear dependence on the covariates *βX_t_* in eq. (11) further helps to explain observed data. The better this component contributes to the prediction task, the lower the local level component *µ_t_* should be. Finally, 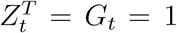 models the noise associated with measuring *y_t_*.

In order to estimate the impact of Governmental interventions, we follow the following methodology. During the training period, we sample the model parameters and the state vector using the Gibbs sampler – a MCMC framework – against the observed data. For the estimation of the impact on the mobility metric, the training period extends over the first 10 days in our selected dataset, i.e, from 10 days prior to the start of Governmental intervention to the start of the intervention. For the estimation of the impact on the effective reproduction number, the training period extends over the first 20 days in our selected dataset, i.e, from 10 days prior and 10 days following the start of Governmental intervention. We select this span of time since, for almost 50% of the people, it takes at least 9.92 days following the date of exposure to get a clinical confirmation of the viral infection (see fig. 10 for the Hessen state in Germany). Hence, on average, there is a delay of around 10 days between the date of exposure and the date of confirmation, and there will not be any immediate effect on the the daily confirmed cases as a result of the intervention. Thus, we define the estimated *effective intervention date* as being equal to the exact intervention date plus 9.92 ≈ 10 days. We then use the posterior simulations to simulate from the posterior predictive distribution over the counterfactual time series given the observed pre-intervention activity during the training. We use the following 30 days for mobility metric and following 20 days for the effective reproduction number for this purpose.

Finally, we use the posterior predictive samples to compute the posterior distribution of the point wise impact, i.e, 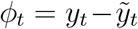, where *y_t_* is the observed quantity and 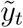 is the counterfactual prediction assuming no intervention. We define the absolute impact of the intervention as the expected value of the point wise impact i.e, *I* = 〈*ϕ*_*t*_〉_*t*_. We use the 30 days and 20 days of posterior predictive samples for mobility metric and *R_t_* to estimate the point wise impact as well as absolute impact because of the intervention. We also estimate the p-value of the observe absolute impact, which measures the probability of obtaining the impact by chance under the null of no intervention.

## Data Availability

Datasets are publicly available.

## Acknowledgments

We thank Ananya Acharya, Dmitry Chernov, Didier Darcet, Euan Mearns, Shyam Nandan, Michael Schatz, Adriana Schellenbaum-Lenner and Ke Wu, for many enlightening discussions during the preparation of the manuscript. Authors contributions: S.R. and D.S. contributed to the design and implementation of the research, to the analysis of the results and to the writing of the manuscript. Competing interests: The authors declare no competing interests. Data and materials availability: All data, code, and materials used in the analysis would be made available after the publication.

## Supplementary Materials

### 5 Supporting figures

**Figure S1:**
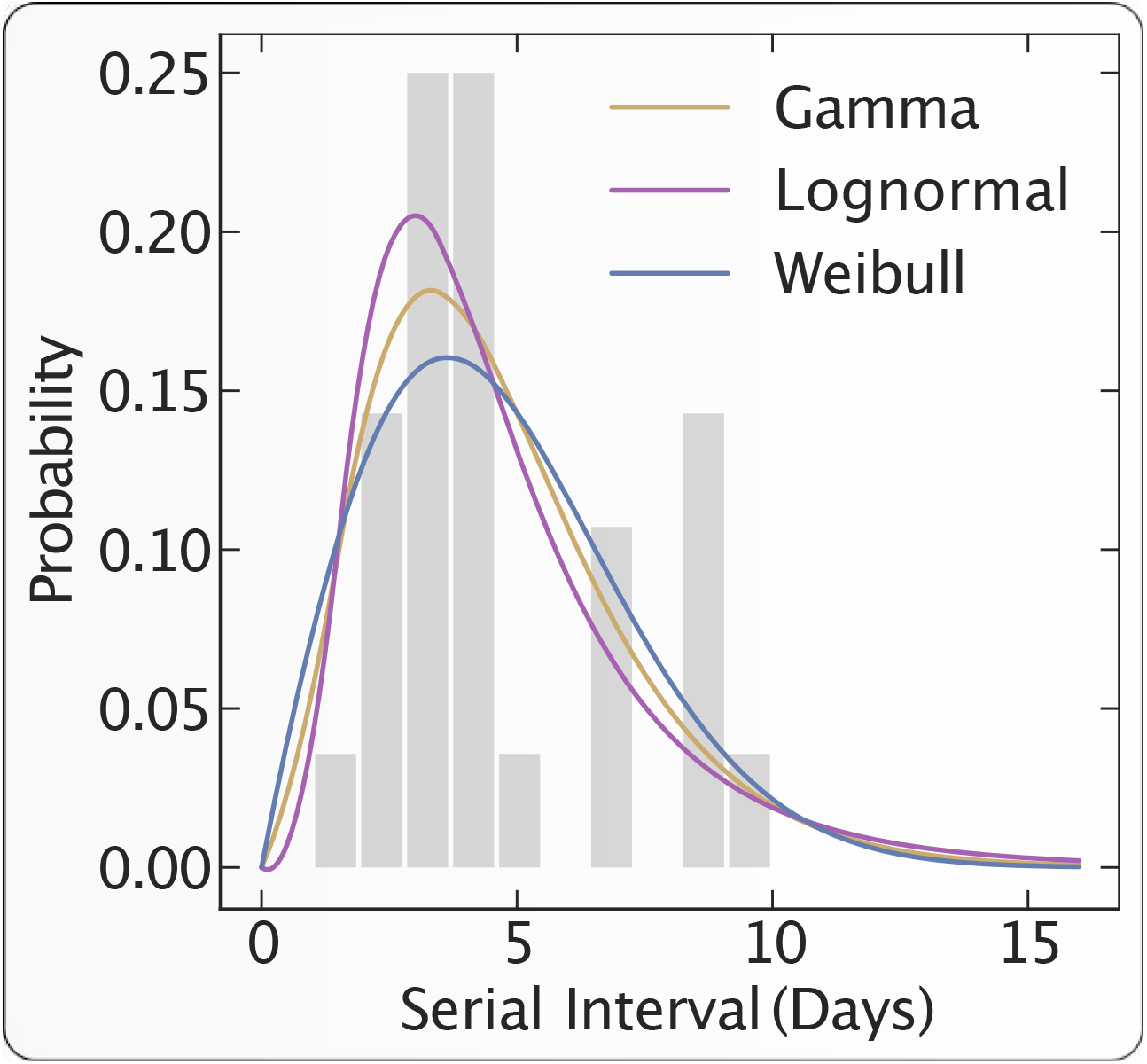
Generation time distribution. The distribution of the serial intervals are calibrated with 3 different models, i.e, Gamma, Log-normal and Weibull and using maximum likelihood estimation. We find the most suitable model for the generation time distribution by comparing the log-likelihood scores of the above models. We observe that the log-normal distribution gives the best fitting with *μ* = 4.6 and *σ* = 2.8. The grey bars represents the empirical observations and the three fitted models are presented as solid lines.

### 6 Supporting Tables

#### 6.1 Impact of interventions on effective reproduction number and growth of epidemic

**Table S1:**
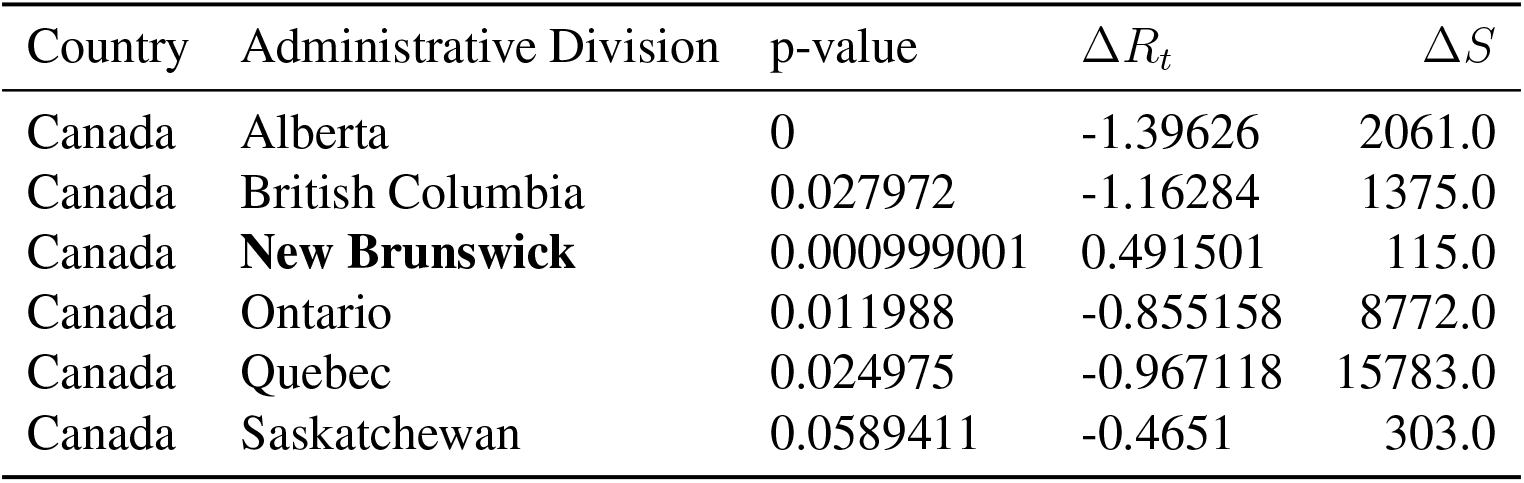
Impact of intervention on *R_t_* and the total growth of confirmed cases in Canada

**Table S2:**
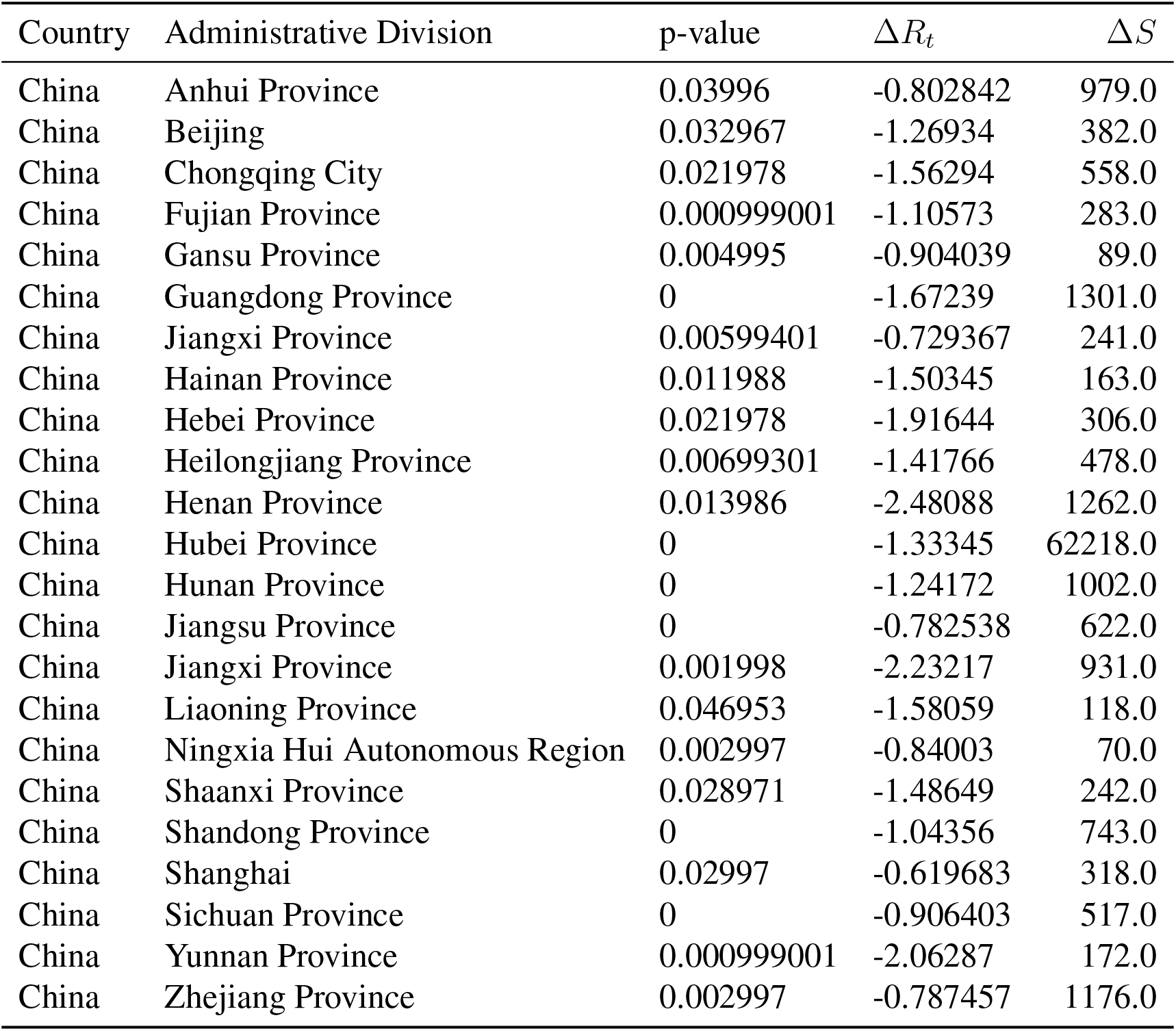
Impact of intervention on *R_t_* and the total growth of confirmed cases in China

| Country | Administrative Division | p-value | $\Delta R_t$ | $\Delta S$ |
| --- | --- | --- | --- | --- |
| China | Anhui Province | 0.03996 | -0.802842 | 979.0 |
| China | Beijing | 0.032967 | -1.26934 | 382.0 |
| China | Chongqing City | 0.021978 | -1.56294 | 558.0 |
| China | Fujian Province | 0.000999001 | -1.10573 | 283.0 |
| China | Gansu Province | 0.004995 | -0.904039 | 89.0 |
| China | Guangdong Province | 0 | -1.67239 | 1301.0 |
| China | Jiangxi Province | 0.00599401 | -0.729367 | 241.0 |
| China | Hainan Province | 0.011988 | -1.50345 | 163.0 |
| China | Hebei Province | 0.021978 | -1.91644 | 306.0 |
| China | Heilongjiang Province | 0.00699301 | -1.41766 | 478.0 |
| China | Henan Province | 0.013986 | -2.48088 | 1262.0 |
| China | Hubei Province | 0 | -1.33345 | 62218.0 |
| China | Hunan Province | 0 | -1.24172 | 1002.0 |
| China | Jiangsu Province | 0 | -0.782538 | 622.0 |
| China | Jiangxi Province | 0.001998 | -2.23217 | 931.0 |
| China | Liaoning Province | 0.046953 | -1.58059 | 118.0 |
| China | Ningxia Hui Autonomous Region | 0.002997 | -0.84003 | 70.0 |
| China | Shaanxi Province | 0.028971 | -1.48649 | 242.0 |
| China | Shandong Province | 0 | -1.04356 | 743.0 |
| China | Shanghai | 0.02997 | -0.619683 | 318.0 |
| China | Sichuan Province | 0 | -0.906403 | 517.0 |
| China | Yunnan Province | 0.000999001 | -2.06287 | 172.0 |
| China | Zhejiang Province | 0.002997 | -0.787457 | 1176.0 |

**Table S3:**
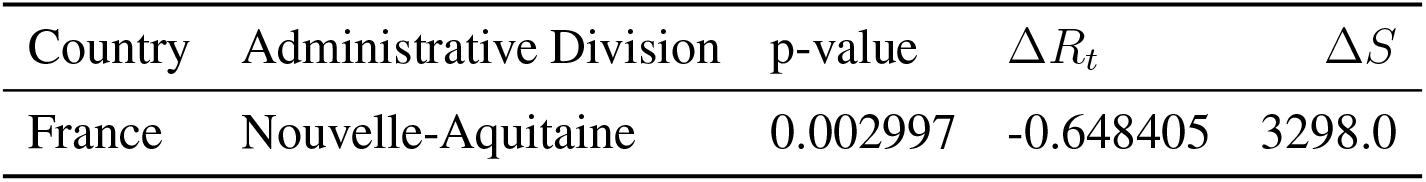
Impact of intervention on *R_t_* and the total growth of confirmed cases in France

**Table S4:**
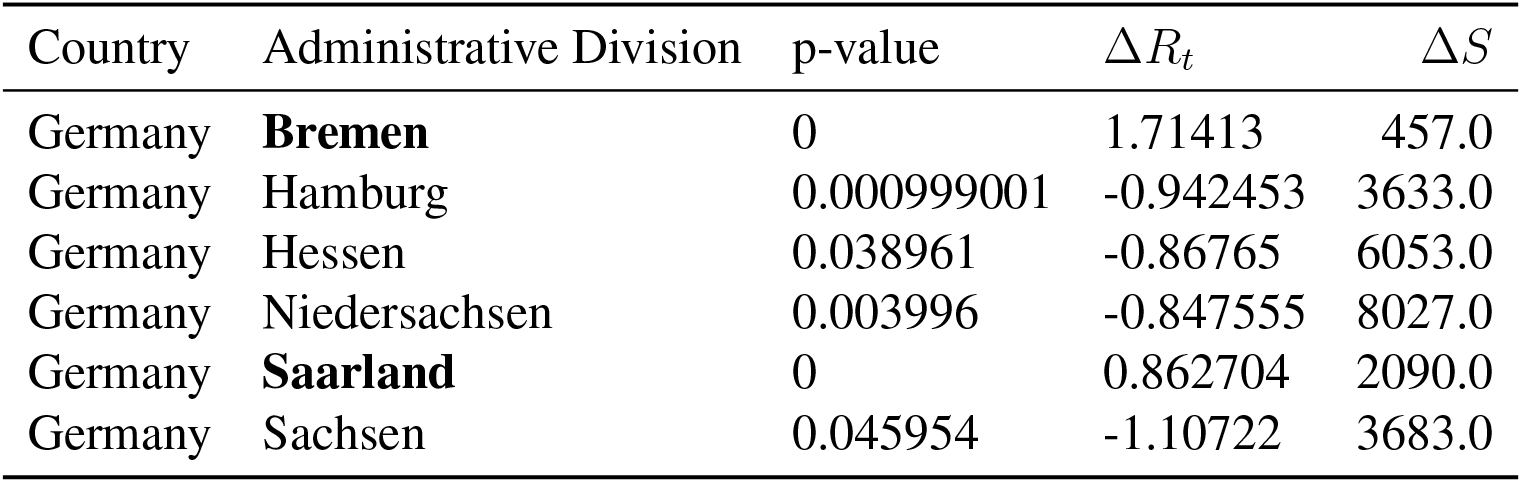
Impact of intervention on *R_t_* and the total growth of confirmed cases in Germany

| Country | Administrative Division | p-value | $\Delta R_t$ | $\Delta S$ |
| --- | --- | --- | --- | --- |
| Germany | **Bremen** | 0 | 1.71413 | 457.0 |
| Germany | Hamburg | 0.000999001 | -0.942453 | 3633.0 |
| Germany | Hessen | 0.038961 | -0.86765 | 6053.0 |
| Germany | Niedersachsen | 0.003996 | -0.847555 | 8027.0 |
| Germany | **Saarland** | 0 | 0.862704 | 2090.0 |
| Germany | Sachsen | 0.045954 | -1.10722 | 3683.0 |

**Table S5:** Impact of intervention on *R_t_* and the total growth of confirmed cases in India

| Country | Administrative Division | p-value | $\Delta R_t$ | $\Delta S$ |
| --- | --- | --- | --- | --- |
| India | **Maharashtra** | 0.002997 | 0.650857 | 6305.0 |
| India | **Odisha** | 0 | 0.494264 | 87.0 |

**Table S6:** Impact of intervention on *R_t_* and the total growth of confirmed cases in Italy

| Country | Administrative Division | p-value | $\Delta R_t$ | $\Delta S$ |
| --- | --- | --- | --- | --- |
| Italy | Puglia | 0.026973 | -0.778201 | 2464.0 |
| Italy | Emilia-Romagna | 0 | -0.381702 | 16439.0 |
| Italy | Liguria | 0.037962 | -0.734625 | 4648.0 |
| Italy | Marche | 0 | -1.00811 | 4387.0 |

**Table S7:** Impact of intervention on *R_t_* and the total growth of confirmed cases in Japan

| Country | Administrative Division | p-value | $\Delta R_t$ | $\Delta S$ |
| --- | --- | --- | --- | --- |
| Japan | Aichi Ken | 0.004995 | -0.553495 | 238.0 |
| Japan | Akita Ken | 0.03996 | -0.362313 | 5.0 |
| Japan | **Chiba Ken** | 0.002997 | 0.297088 | 572.0 |
| Japan | Fukui Ken | 0.000999001 | -0.988835 | 57.0 |
| Japan | Fukuoka Ken | 0.034965 | -0.573188 | 451.0 |
| Japan | Gifu Ken | 0 | -1.02953 | 81.0 |
| Japan | Hyogo Ken | 0.00799201 | -0.721471 | 447.0 |
| Japan | Ibaraki Ken | 0.048951 | -0.935754 | 90.0 |
| Japan | Kochi Ken | 0.017982 | -0.676786 | 36.0 |
| Japan | Kyoto Fu | 0.00799201 | -0.416234 | 196.0 |
| Japan | Miyagi Ken | 0 | -1.02827 | 56.0 |
| Japan | Okayama Ken | 0.00899101 | -0.567645 | 11.0 |
| Japan | Osaka Fu | 0.016983 | -0.359852 | 1217.0 |
| Japan | **Saga Ken** | 0 | 0.564563 | 34.0 |
| Japan | Tochigi Ken | 0.0579421 | -0.567938 | 34.0 |
| Japan | Tokyo To | 0.036963 | -0.201202 | 3611.0 |
| Japan | Yamagata Ken | 0.01998 | -0.612601 | 50.0 |
| Japan | Yamaguchi Ken | 0.037962 | -0.4729 | 21.0 |
| Japan | Yamanashi Ken | 0.00999001 | -0.855139 | 34.0 |

**Table S8:** Impact of intervention on *R_t_* and the total growth of confirmed cases in Spain

| Country | Administrative Division | p-value | $\Delta R_t$ | $\Delta S$ |
| --- | --- | --- | --- | --- |
| Spain | Asturias | 0.048951 | -0.571728 | 1958.0 |
| Spain | La Rioja | 0.00899101 | -0.562582 | 3304.0 |
| Spain | Madrid | 0.000999001 | -0.568361 | 48108.0 |

**Table S9:** Impact of intervention on *R_t_* and the total growth of confirmed cases in Switzerland

| Country | Administrative Division | p-value | $\Delta R_t$ | $\Delta S$ |
| --- | --- | --- | --- | --- |
| Switzerland | Appenzell Ausserrhoden | 0.04995 | -2.53811 | 68.0 |
| Switzerland | Bern | 0.016983 | -1.3457 | 1347.0 |
| Switzerland | Fribourg | 0 | -0.764614 | 834.0 |
| Switzerland | Genève | 0 | -0.749412 | 3902.0 |
| Switzerland | Graubünden | 0.0599401 | -0.690593 | 625.0 |
| Switzerland | **Luzern** | 0 | 0.384942 | 524.0 |
| Switzerland | Neuchâtel | 0.001998 | -0.370057 | 506.0 |
| Switzerland | **Solothurn** | 0 | 0.292663 | 282.0 |
| Switzerland | St. Gallen | 0.0569431 | -2.59314 | 617.0 |
| Switzerland | Ticino | 0.00899101 | -1.08246 | 2544.0 |
| Switzerland | Valais | 0.001998 | -1.13367 | 1535.0 |
| Switzerland | Vaud | 0.00599401 | -1.08239 | 4125.0 |
| Switzerland | Zürich | 0 | -1.19565 | 2739.0 |

**Table S10:** Impact of intervention on *R_t_* and the total growth of confirmed cases in United States

| Country | Administrative Division | p-value | $\Delta R_t$ | $\Delta S$ |
| --- | --- | --- | --- | --- |
| United States | **Alaska** | 0.0559441 | 0.44278 | 303.0 |
| United States | Colorado | 0.00599401 | -1.53705 | 8459.0 |
| United States | Connecticut | 0.004995 | -1.7927 | 16713.0 |
| United States | Florida | 0.004995 | -1.27691 | 23729.0 |
| United States | Georgia | 0 | -0.887848 | 16907.0 |
| United States | Louisiana | 0.000999001 | -1.28319 | 22771.0 |
| United States | Maryland | 0.002997 | -0.52522 | 11465.0 |
| United States | Massachusetts | 0 | -0.700434 | 34896.0 |
| United States | Michigan | 0 | -0.460859 | 26631.0 |
| United States | Minnesota | 0 | -0.768463 | 1982.0 |
| United States | Mississippi | 0.035964 | -0.80443 | 3743.0 |
| United States | New Jersey | 0.040959 | -0.934117 | 77725.0 |
| United States | Pennsylvania | 0.011988 | -0.584873 | 29256.0 |
| United States | South Carolina | 0.03996 | -1.14461 | 3871.0 |
| United States | **South Dakota** | 0 | 1.08317 | 1400.0 |
| United States | **Wyoming** | 0.020979 | 0.536284 | 278.0 |

#### 6.2 Names of different administrative divisions and their ISO code

**Table S11:** Different administrative divisions and corresponding ISO code

| Country | Name | Code |
| --- | --- | --- |
| Italy | Piemonte | 21 |
| Italy | Valle d'Aosta/Valle d'Aoste | 23 |
| Italy | Lombardia | 25 |
| Italy | Trentino-Alto Adige/Sdtirol | 32 |
| Italy | Veneto | 34 |
| Italy | Friuli-Venezia Giulia | 36 |
| Italy | Liguria | 42 |
| Italy | Emilia-Romagna | 45 |
| Italy | Toscana | 52 |
| Italy | Umbria | 55 |
| Italy | Marche | 57 |
| Italy | Lazio | 62 |
| Italy | Abruzzo | 65 |
| Italy | Molise | 67 |
| Italy | Campania | 72 |
| Italy | Puglia | 75 |
| Italy | Basilicata | 77 |
| Italy | Calabria | 78 |
| Italy | Sicilia | 82 |
| Italy | Sardegna | 88 |

**Table S12:**
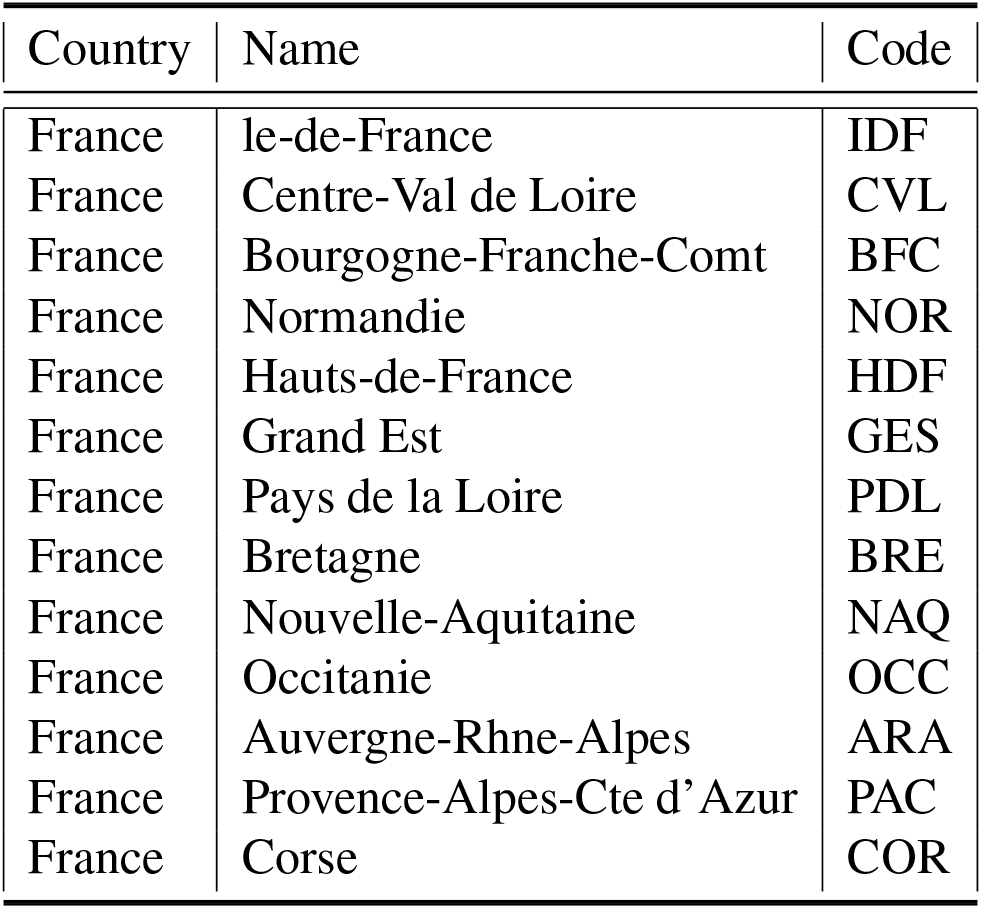
Different administrative divisions and corresponding ISO code

**Table S13:** Different administrative divisions and corresponding ISO code

| Country | Name | Code |
| --- | --- | --- |
| China | Xinjiang Uygur Autonomous Region | 65 |
| China | Tibet Autonomous Region | XZ |
| China | Inner Mongolia Autonomous Region | 15 |
| China | Qinghai Province | 63 |
| China | Sichuan Province | 51 |
| China | Heilongjiang Province | 23 |
| China | Gansu Province | 62 |
| China | Yunnan Province | 53 |
| China | Guangxi Zhuang Autonomous Region | 45 |
| China | Hunan Province | 43 |
| China | Shaanxi Province | 61 |
| China | Guangdong Province | 44 |
| China | Jilin Province | 22 |
| China | Hebei Province | 13 |
| China | Hubei Province | 42 |
| China | Guizhou Province | 52 |
| China | Shandong Province | 37 |
| China | Jiangxi Province | 36 |
| China | Henan Province | 41 |
| China | Liaoning Province | 21 |
| China | Shanxi Province | 14 |
| China | Anhui Province | 34 |
| China | Fujian Province | 35 |
| China | Zhejiang Province | 33 |
| China | Jiangsu Province | 32 |
| China | Chongqing City | 50 |
| China | Ningxia Hui Autonomous Region | 64 |
| China | Hainan Province | 46 |
| China | Taiwan Province | 71 |
| China | Beijing | 11 |
| China | Tianjin City | 12 |
| China | Shanghai | 31 |
| China | Hong Kong Special Administrative Region | 50 |
| China | Macao Special Administrative Region | MO |

**Table S14:** Different administrative divisions and corresponding ISO code

| Country | Name | Code |
| --- | --- | --- |
| Switzerland | Graubnden | GR |
| Switzerland | Bern | BE |
| Switzerland | Valais | VS |
| Switzerland | Vaud | VD |
| Switzerland | Ticino | TI |
| Switzerland | St. Gallen | SG |
| Switzerland | Zrich | ZH |
| Switzerland | Fribourg | FR |
| Switzerland | Luzern | LU |
| Switzerland | Aargau | AG |
| Switzerland | Uri | UR |
| Switzerland | Thurgau | TG |
| Switzerland | Schwyz | SZ |
| Switzerland | Jura | JU |
| Switzerland | Neuchtel | NE |
| Switzerland | Solothurn | SO |
| Switzerland | Glarus | GL |
| Switzerland | Basel-Landschaft | BL |
| Switzerland | Obwalden | OW |
| Switzerland | Nidwalden | NW |
| Switzerland | Genve | GE |
| Switzerland | Schaffhausen | SH |
| Switzerland | Appenzell Ausserrhoden | AR |
| Switzerland | Zug | ZG |
| Switzerland | Appenzell Innerrhoden | AI |
| Switzerland | Basel-Stadt | BS |

**Table S15:** Different administrative divisions and corresponding ISO code

| Country | Name | Code |
| --- | --- | --- |
| Spain | Castilla-Leon | CL |
| Spain | Catalua | CT |
| Spain | Ceuta | CE |
| Spain | Murcia | MU |
| Spain | La Rioja | LO |
| Spain | Baleares | PM |
| Spain | Canarias | CN |
| Spain | Cantabria | CB |
| Spain | Andalucia | AN |
| Spain | Asturias | O |
| Spain | Valencia | P |
| Spain | Melilla | ML |
| Spain | Navarra | NC |
| Spain | Galicia | GA |
| Spain | Aragon | AR |
| Spain | Madrid | M |
| Spain | Extremadura | EX |
| Spain | Castilla-La Mancha | CM |
| Spain | Pais Vasco | PV |

**Table S16:**
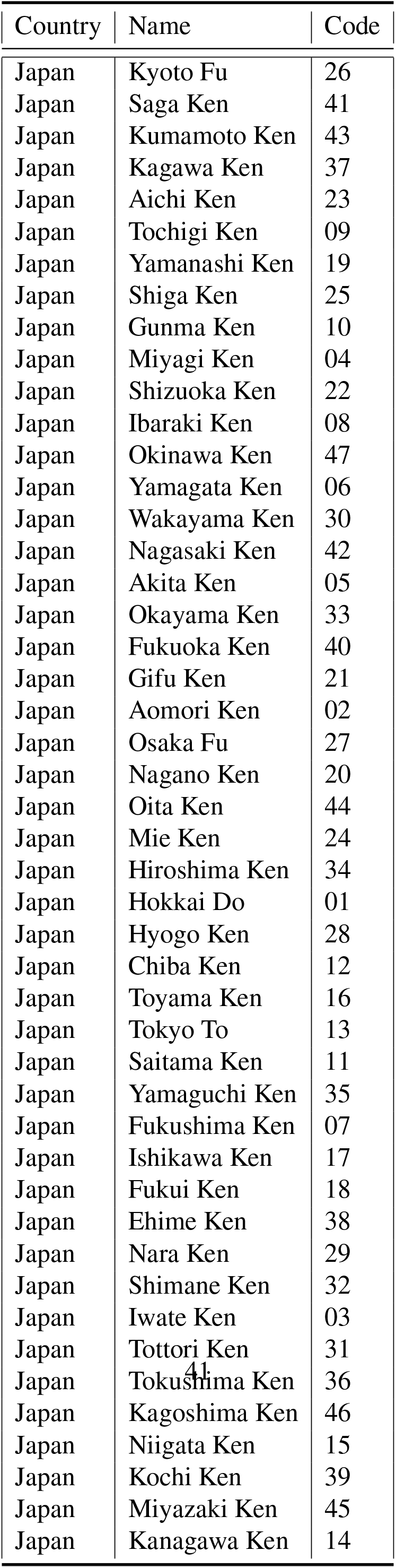
Different administrative divisions and corresponding ISO code.

**Table S17:**
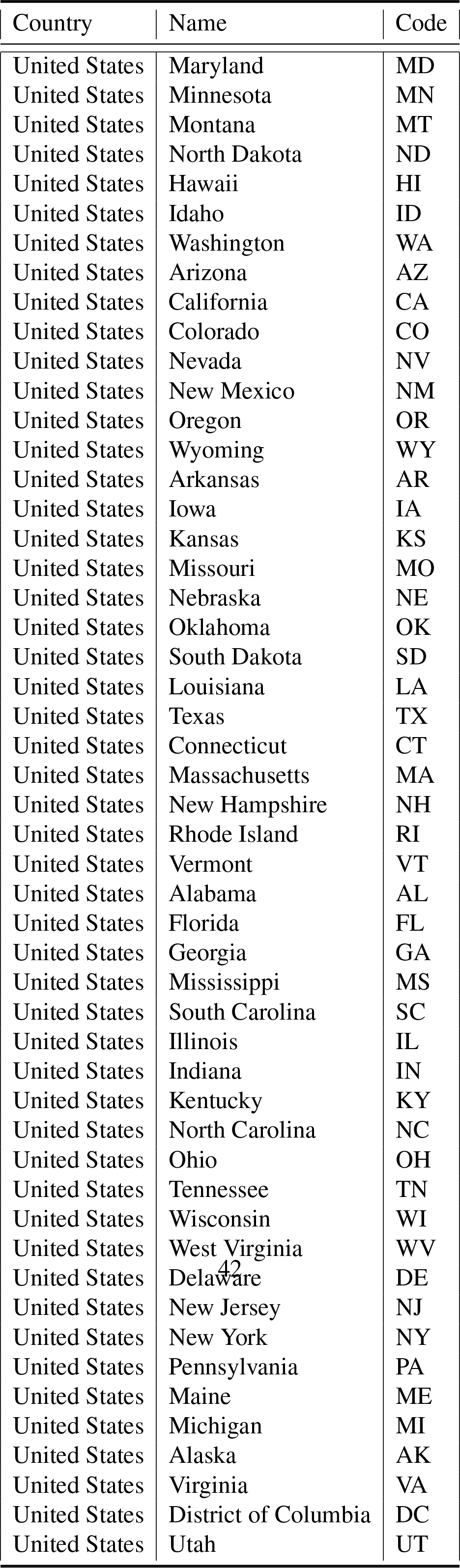
Different administrative divisions and corresponding ISO code.

**Table S18:**
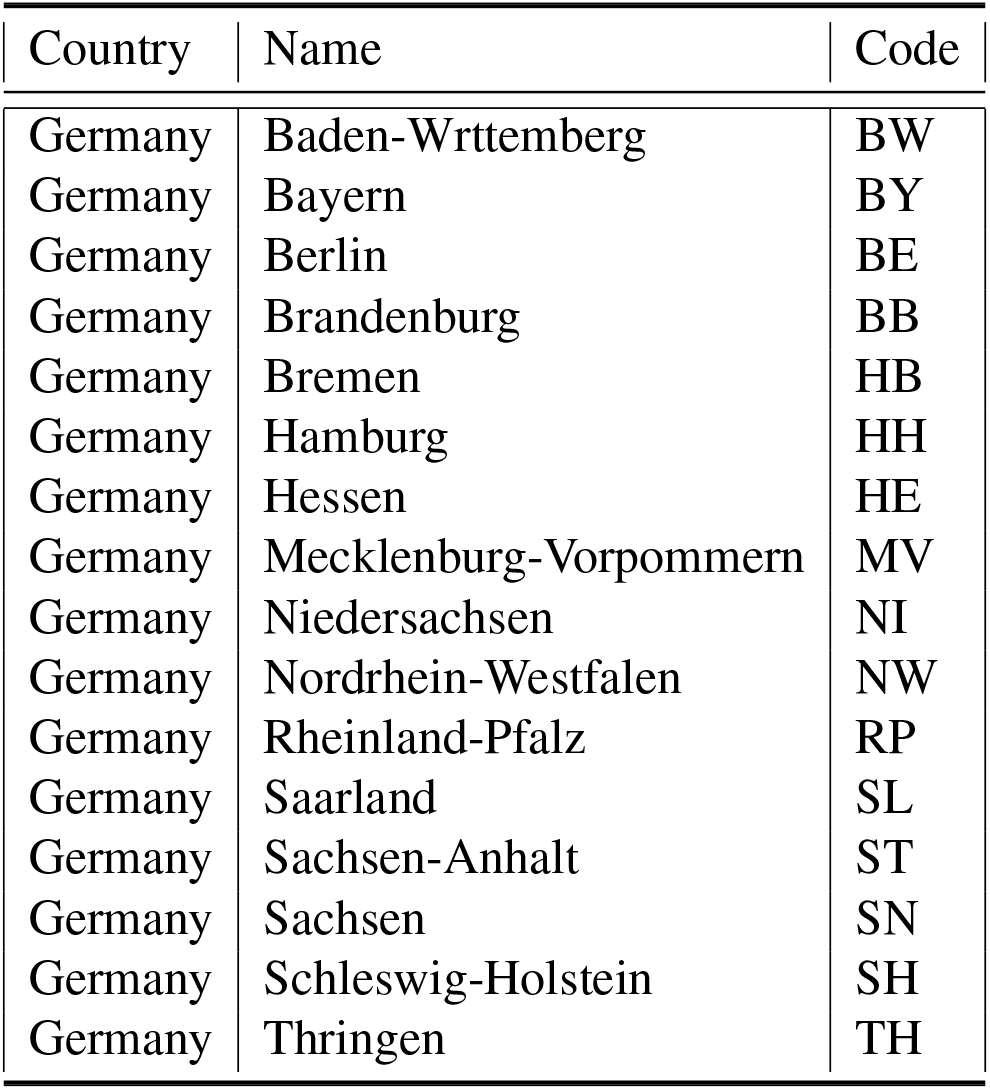
Different administrative divisions and corresponding ISO code

**Table S19:** Different administrative divisions and corresponding ISO code

| Country | Name | Code |
| --- | --- | --- |
| India | Andaman and Nicobar Islands | AN |
| India | Arunachal Pradesh | AR |
| India | Assam | AS |
| India | Bihar | BR |
| India | Chandigarh | CH |
| India | Chhattisgarh | CT |
| India | Dadra and Nagar Haveli | DN |
| India | Daman and Diu | DD |
| India | Goa | GA |
| India | Gujarat | GJ |
| India | Haryana | HR |
| India | Himachal Pradesh | HP |
| India | Jharkhand | JH |
| India | Karnataka | KA |
| India | Kerala | KL |
| India | Lakshadweep | LD |
| India | Madhya Pradesh | MP |
| India | Maharashtra | MH |
| India | Manipur | MN |
| India | Meghalaya | ML |
| India | Mizoram | MZ |
| India | Nagaland | NL |
| India | Delhi | DL |
| India | Puducherry | PY |
| India | Punjab | PB |
| India | Rajasthan | RJ |
| India | Sikkim | SK |
| India | Tamil Nadu | TN |
| India | Telangana | TG |
| India | Tripura | TR |
| India | Uttar Pradesh | UP |
| India | Uttarakhand | UT |
| India | West Bengal | WB |
| India | Odisha | OR |
| India | Andhra Pradesh | AP |
| India | Jammu and Kashmir | JK |
| India | Ladakh | LD |

**Table S20:**
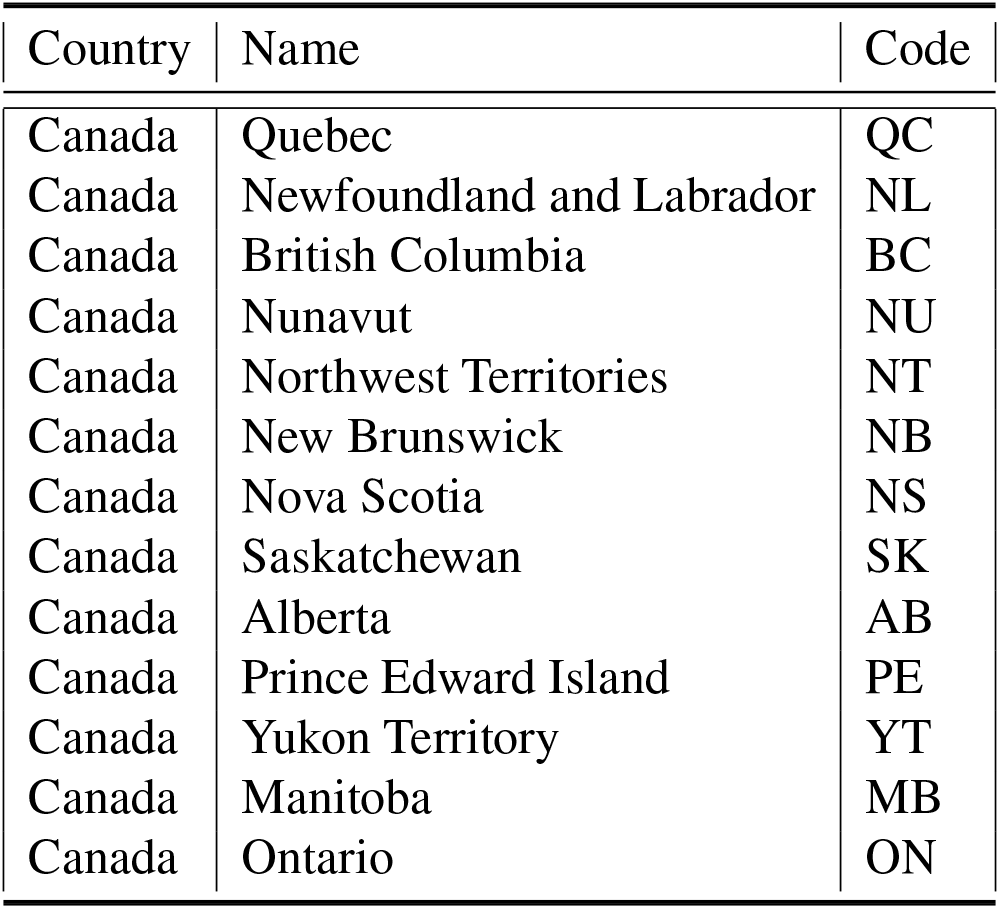
Different administrative divisions and corresponding ISO code

### 7 Supplemental figures

#### 7.1 Mobility Impact

**Figure S2:**
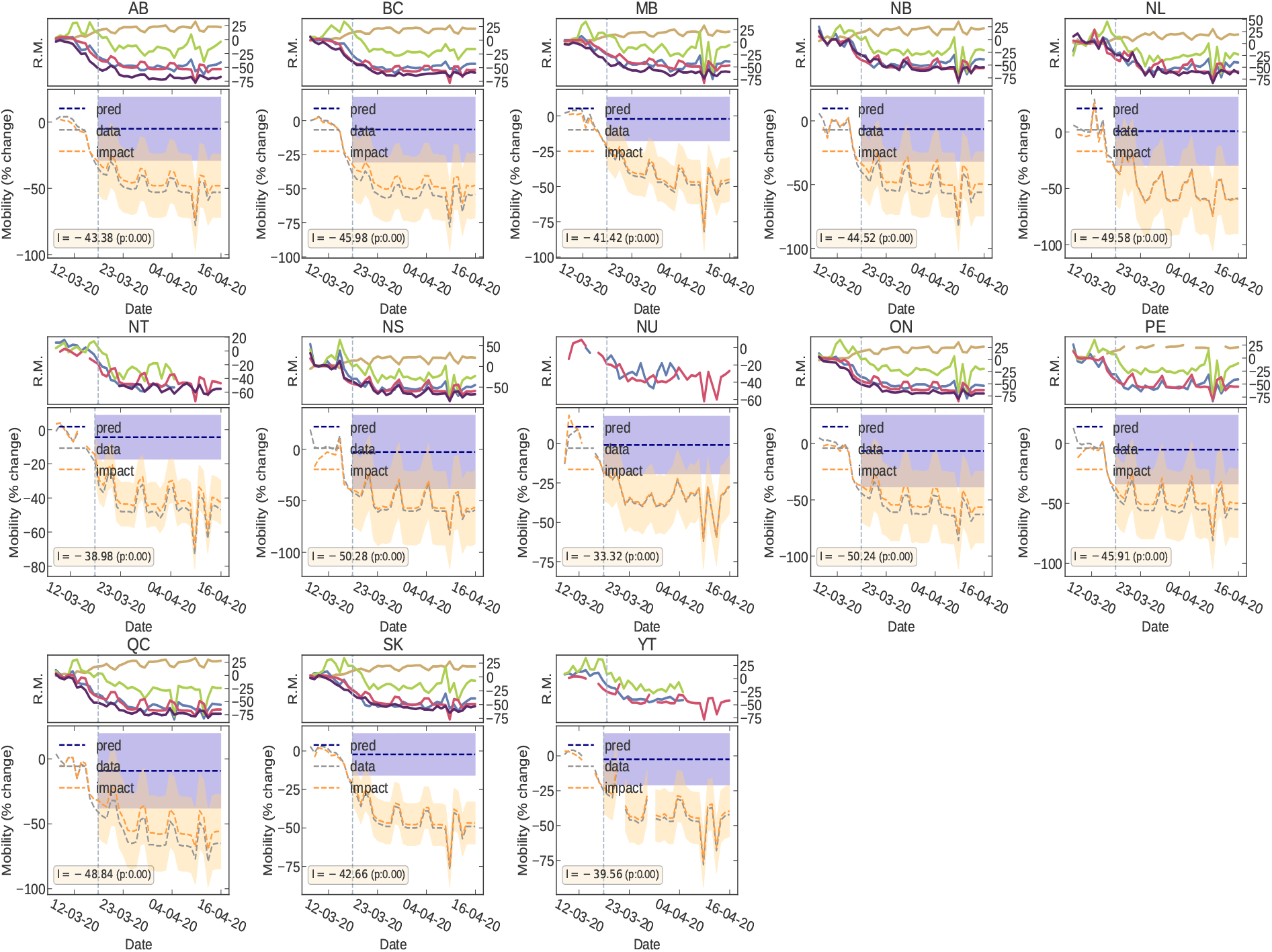
Impact of Governmental interventions on mobility in different administrative divisions of Canada.

**Figure S3:**
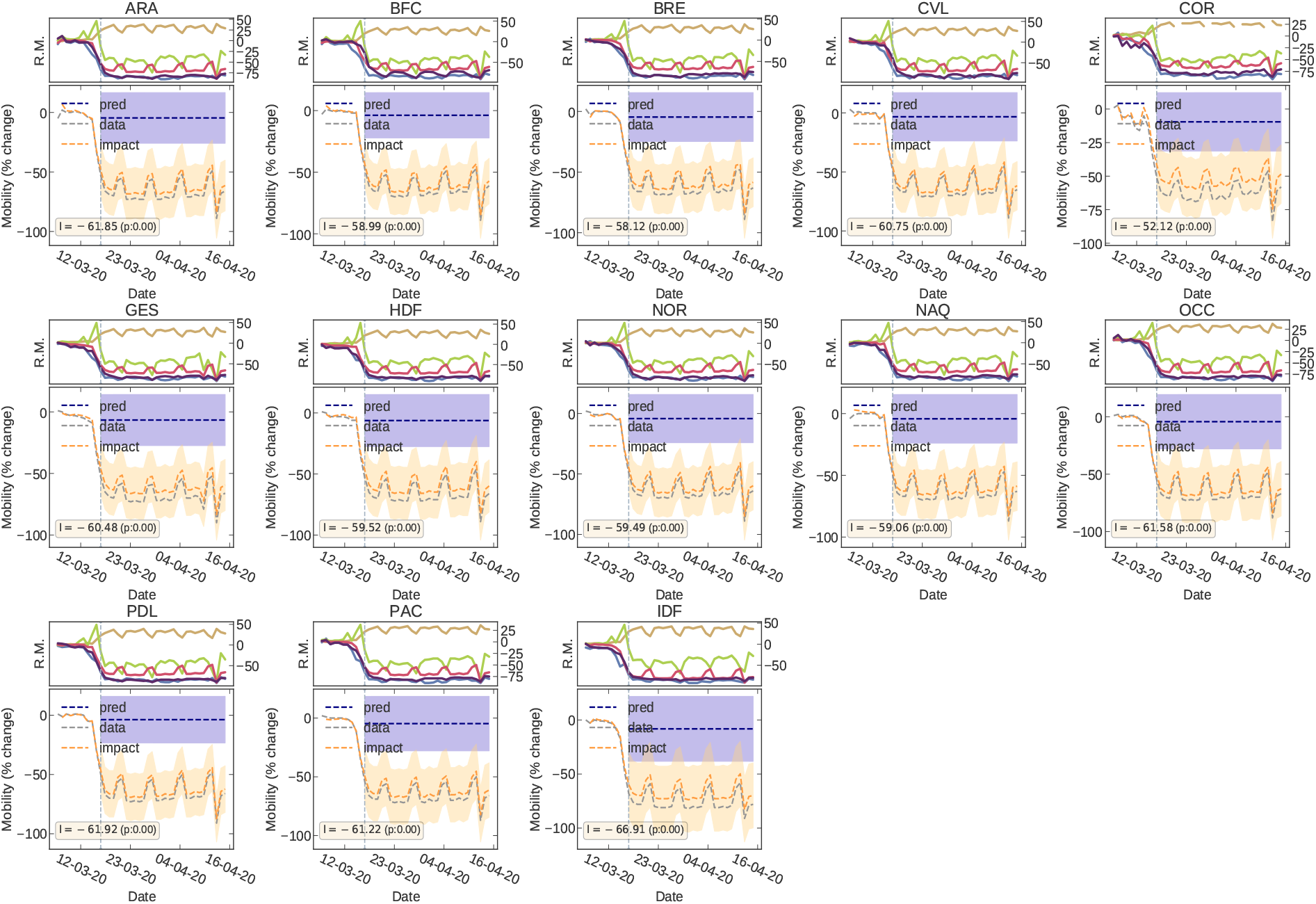
Impact of Governmental interventions on mobility in different administrative divisions of France.

**Figure S4:**
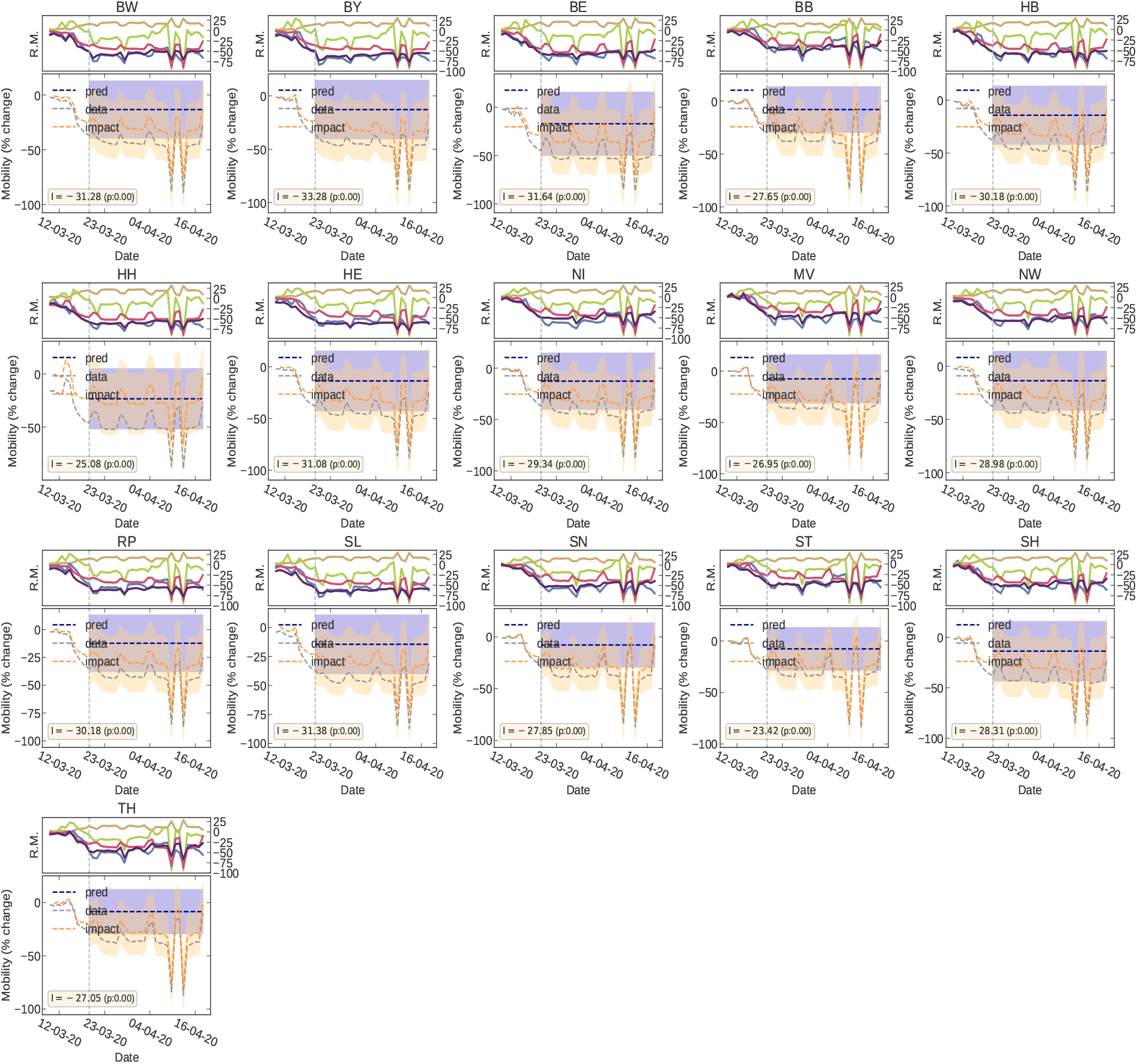
Impact of Governmental interventions on mobility in different administrative divisions of Germany.

**Figure S5:**
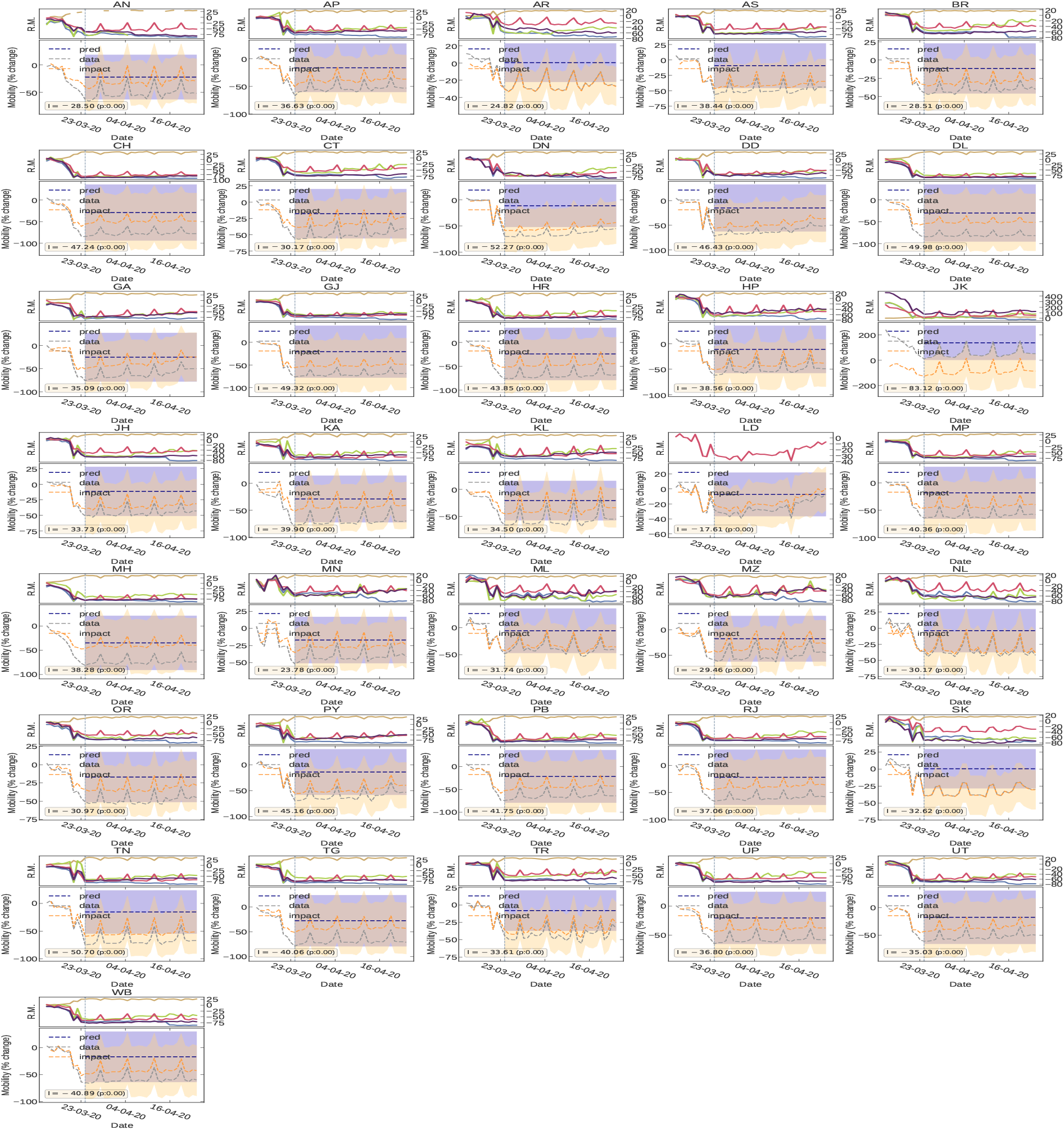
Impact of Governmental interventions on mobility in different administrative divisions of India.

**Figure S6:**
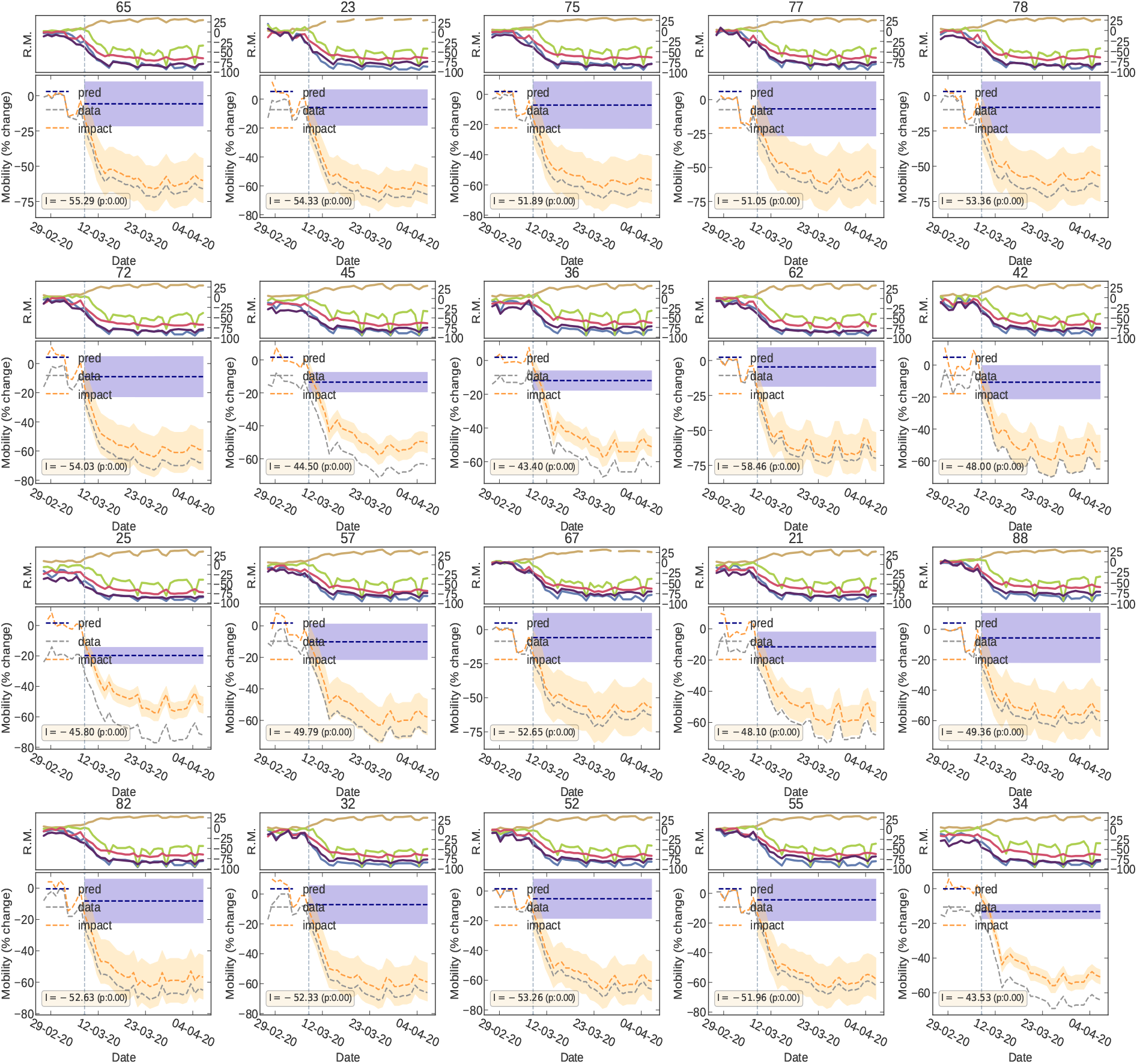
Impact of Governmental interventions on mobility in different administrative divisions of Italy.

**Figure S7:**
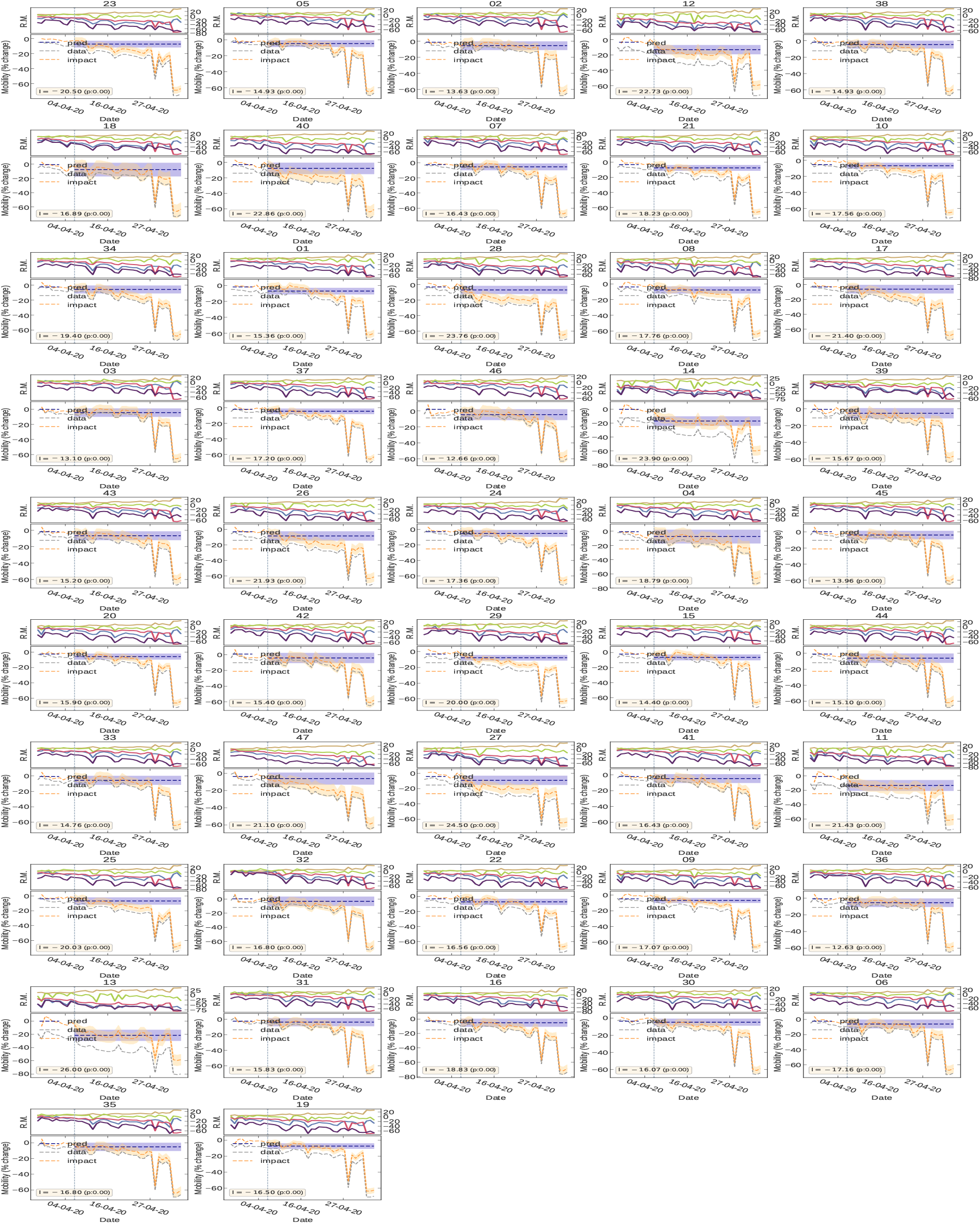
Impact of Governmental interventions on mobility in different administrative divisions of Japan.

**Figure S8:**
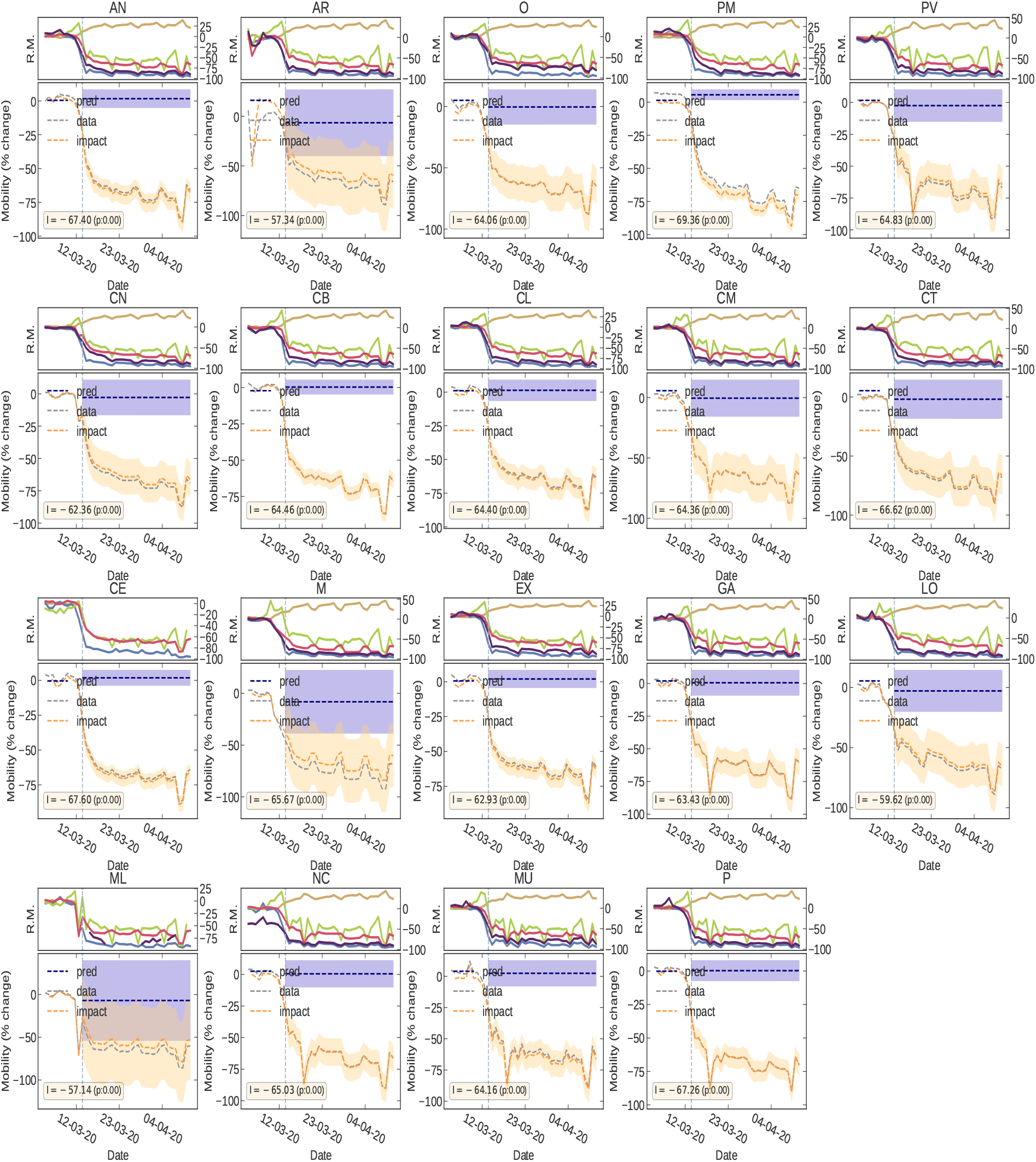
Impact of Governmental interventions on mobility in different administrative divisions of Spain.

**Figure S9:**
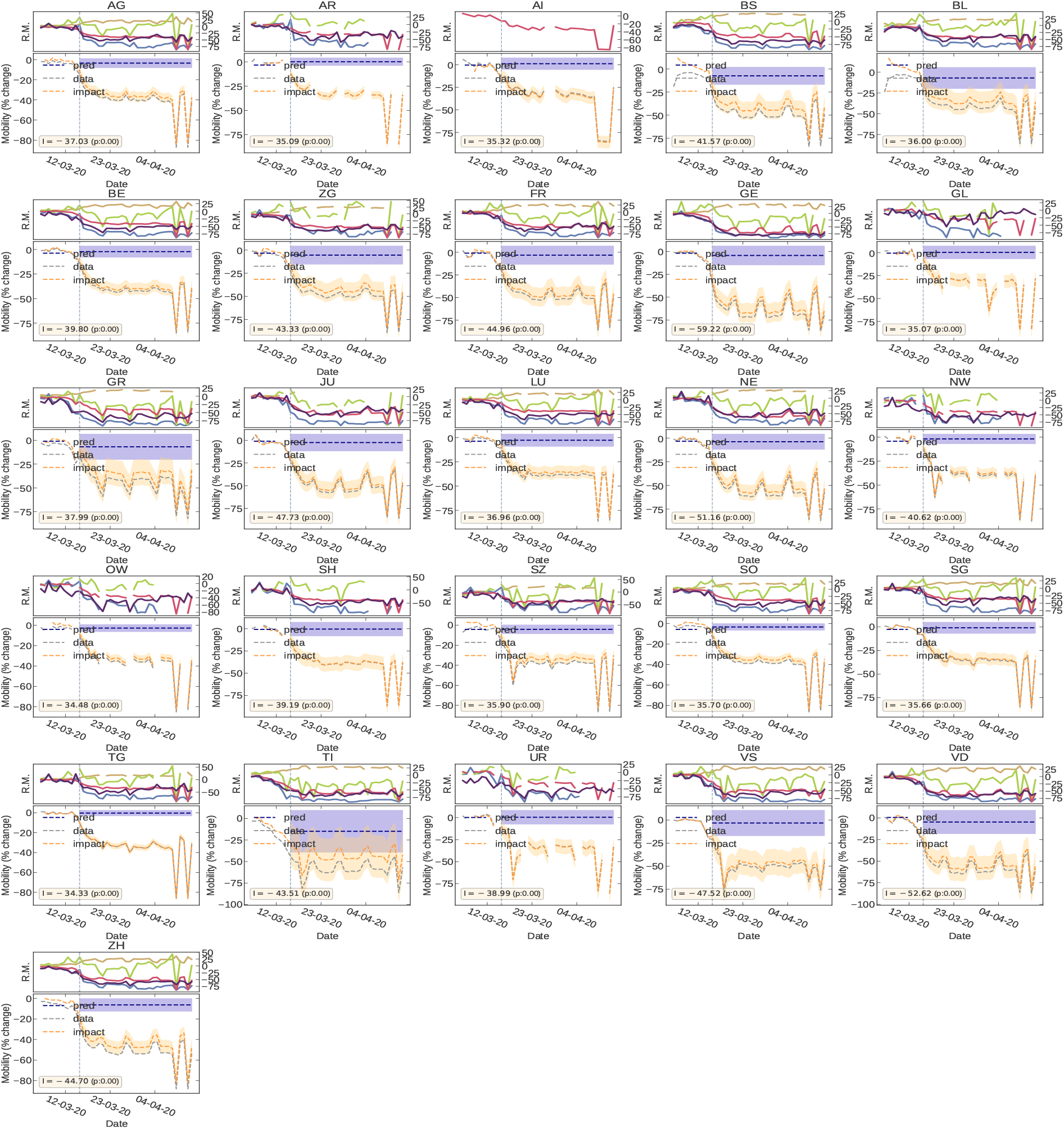
Impact of Governmental interventions on mobility in different administrative divisions of Switzerland.

**Figure S10:**
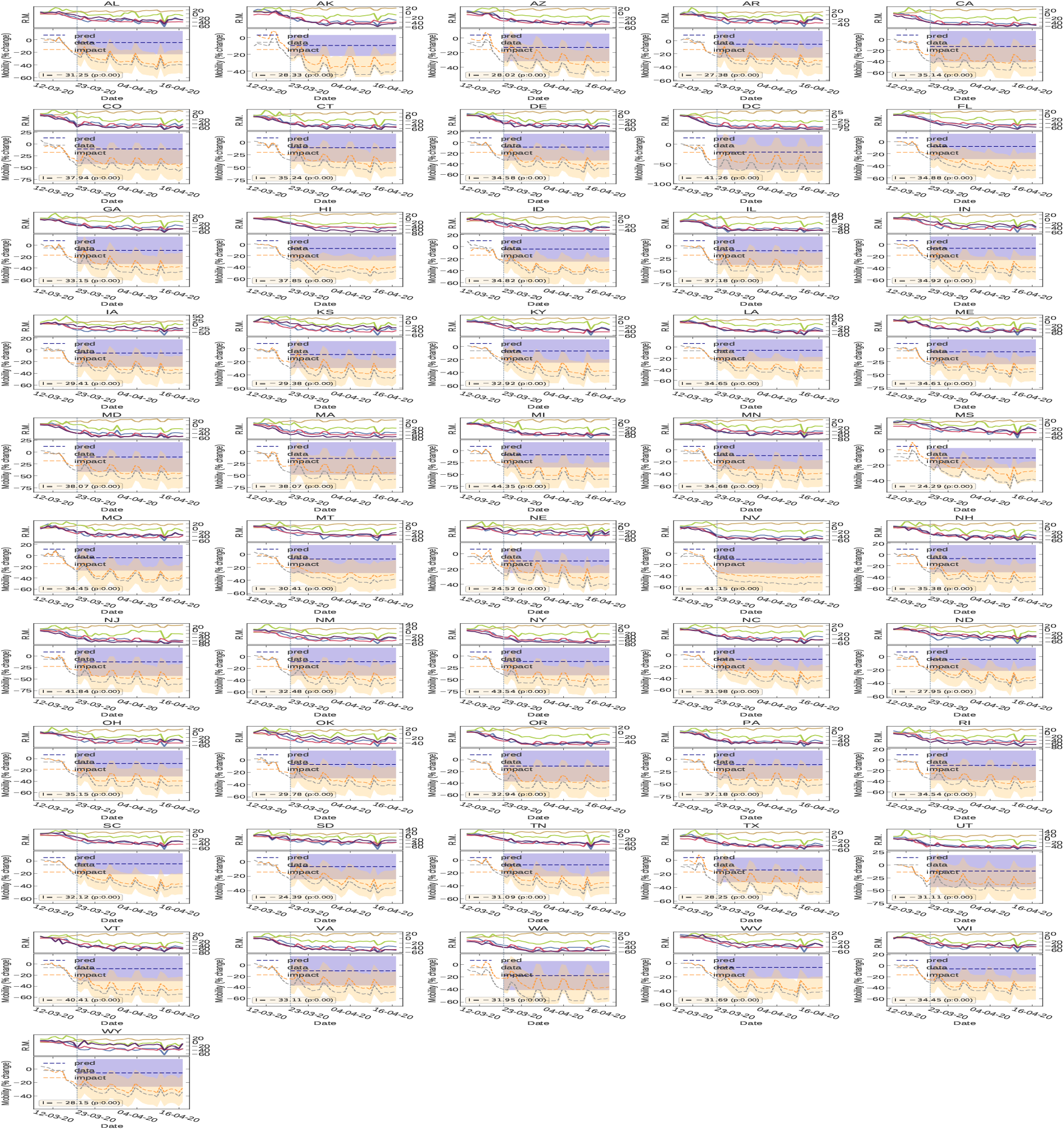
Impact of Governmental interventions on mobility in different administrative divisions of United States.

#### 7.2 Epidemic Impact

**Figure S11:**
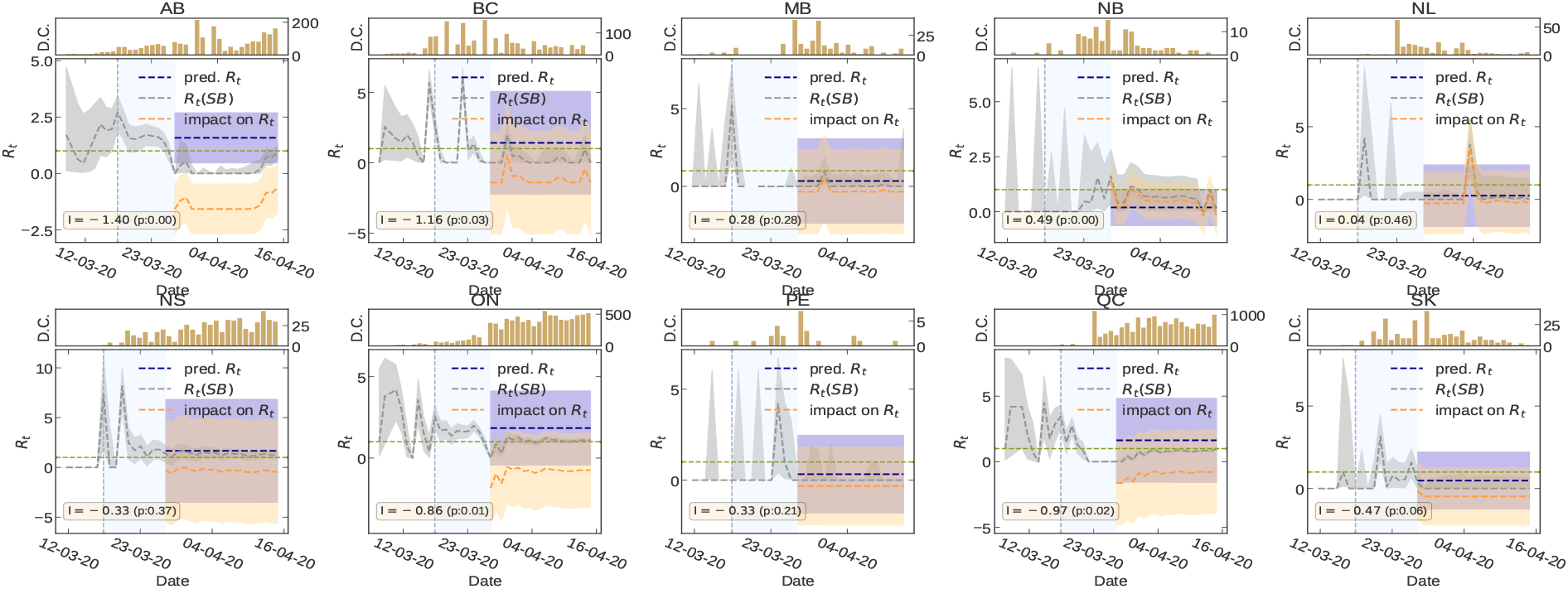
Impact of Governmental interventions on *R_t_* in different administrative divisions of Canada.

**Figure S12:**
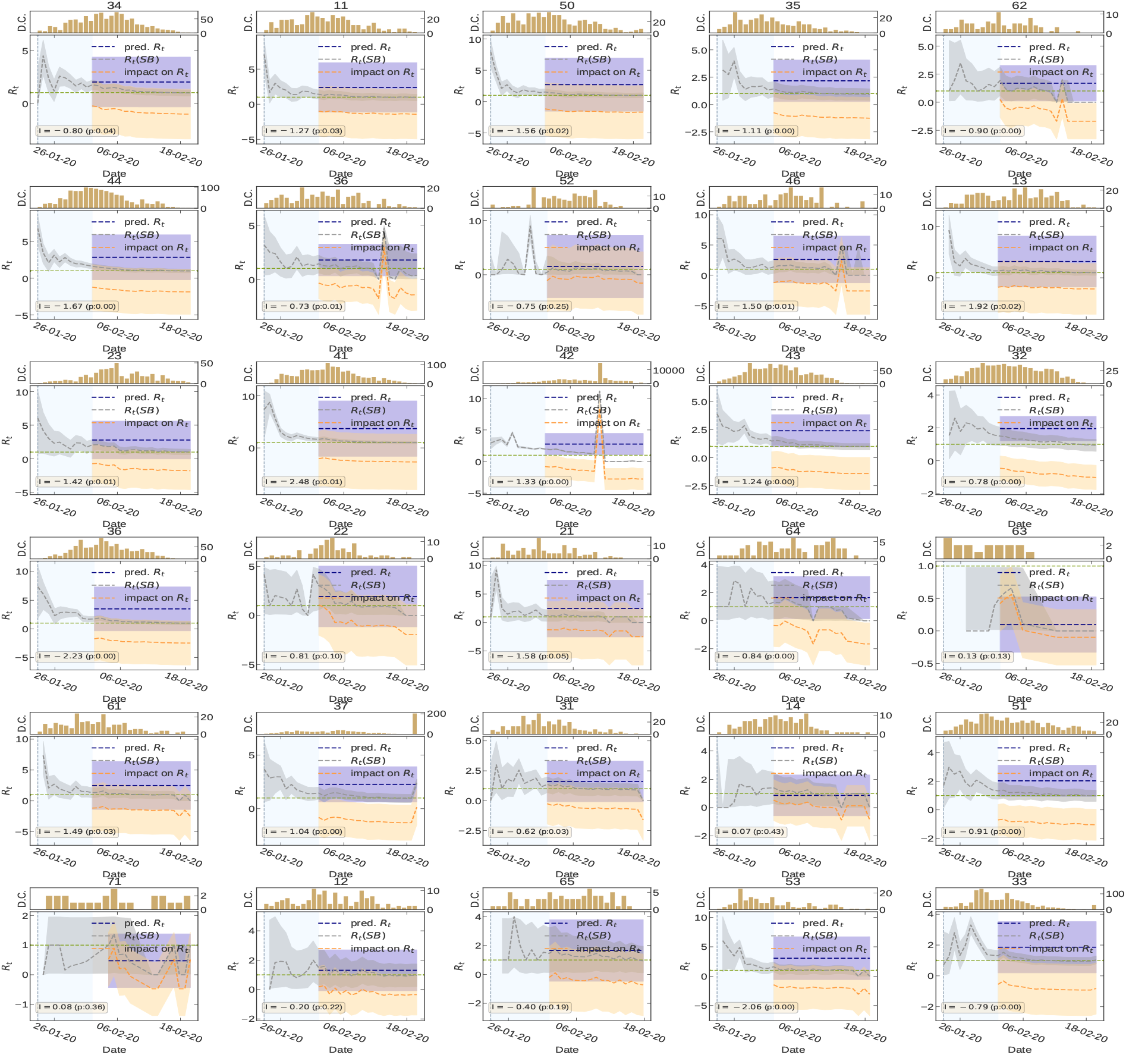
Impact of Governmental interventions on *R_t_* in different administrative divisions of China.

**Figure S13:**
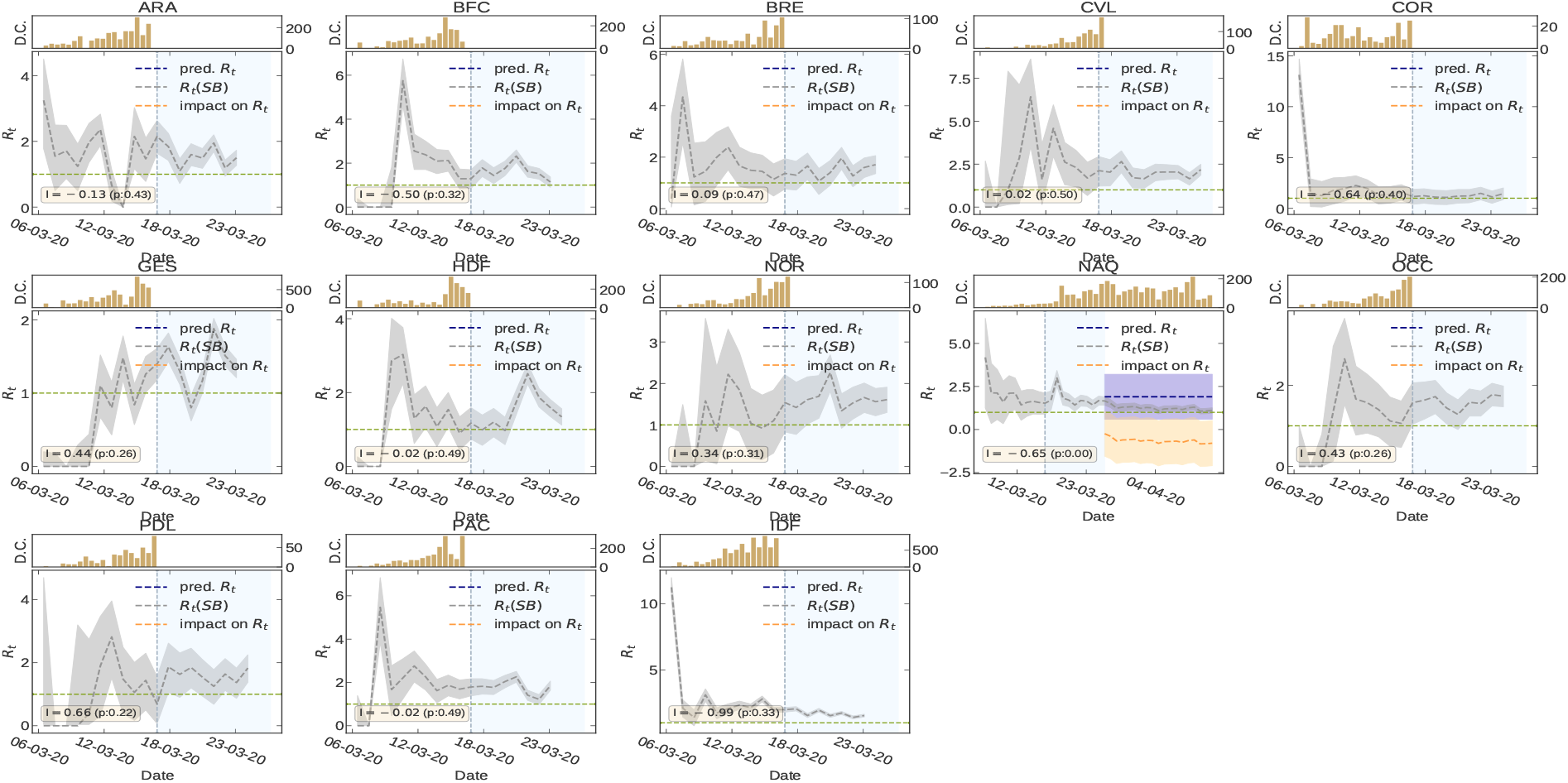
Impact of Governmental interventions on *R_t_* in different administrative divisions of France.

**Figure S14:**
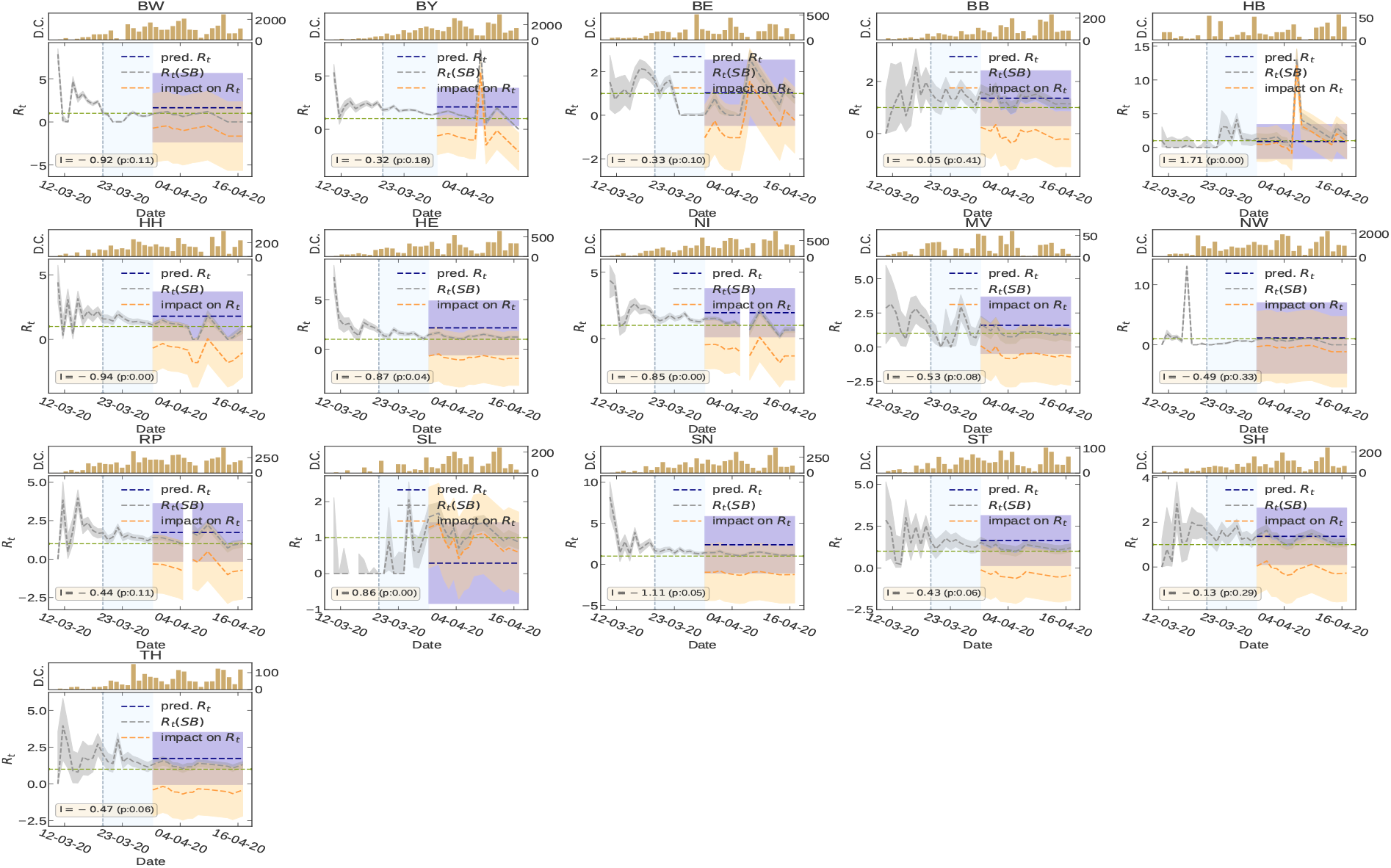
Impact of Governmental interventions on *R_t_* in different administrative divisions of Germany.

**Figure S15:**
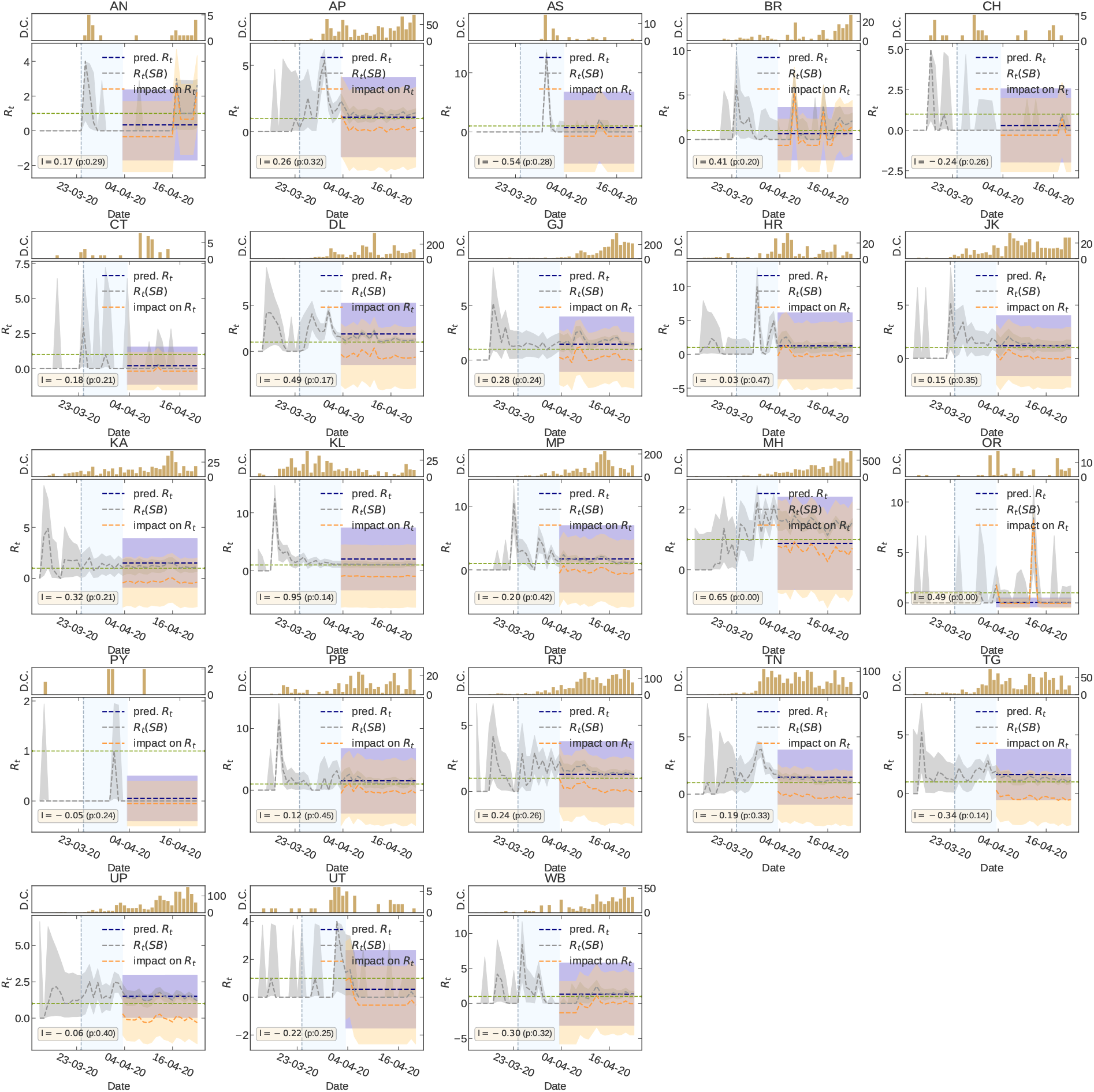
Impact of Governmental interventions on *R_t_* in different administrative divisions of India.

**Figure S16:**
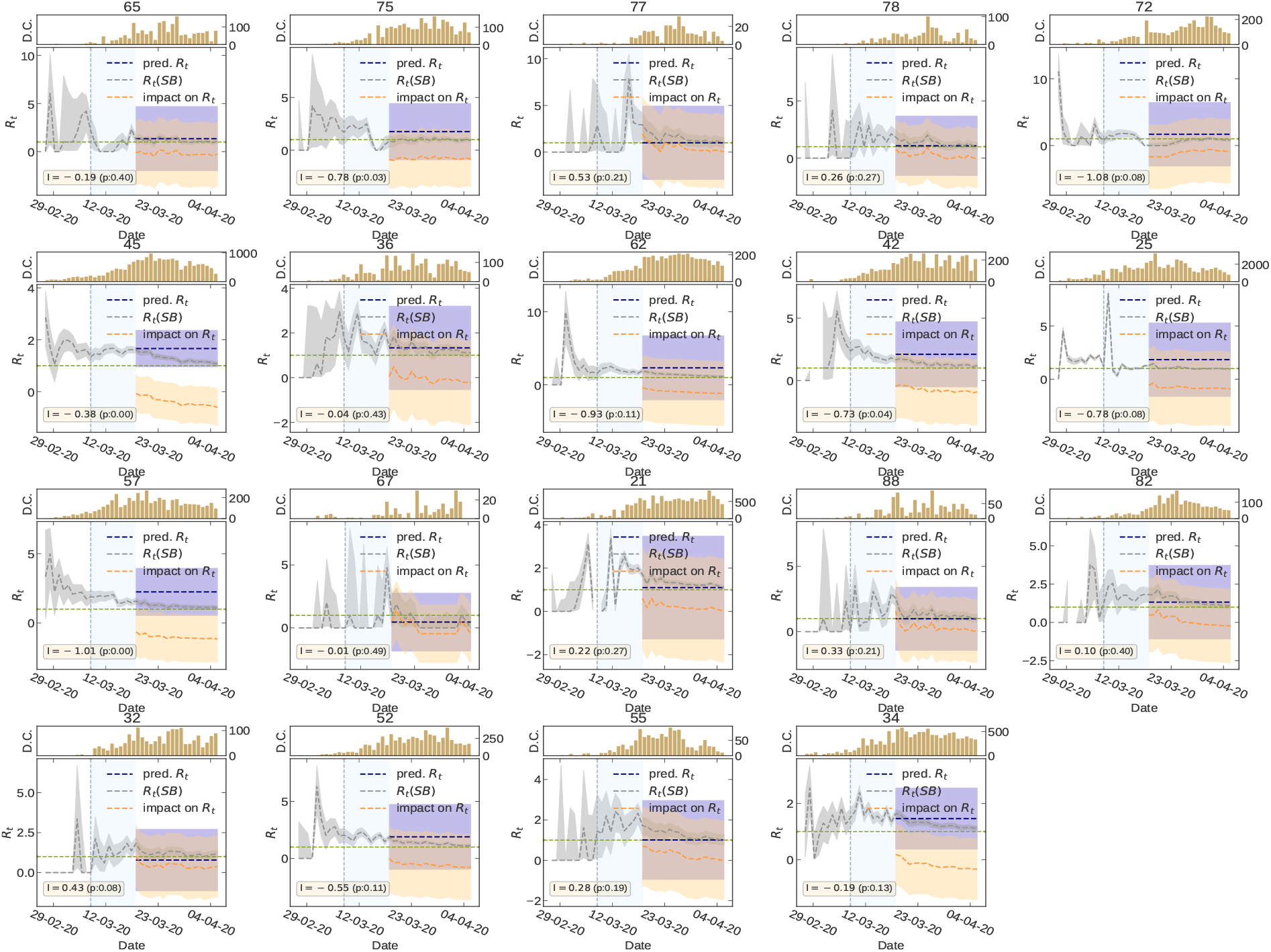
Impact of Governmental interventions on *R_t_* in different administrative divisions of Italy.

**Figure S17:**
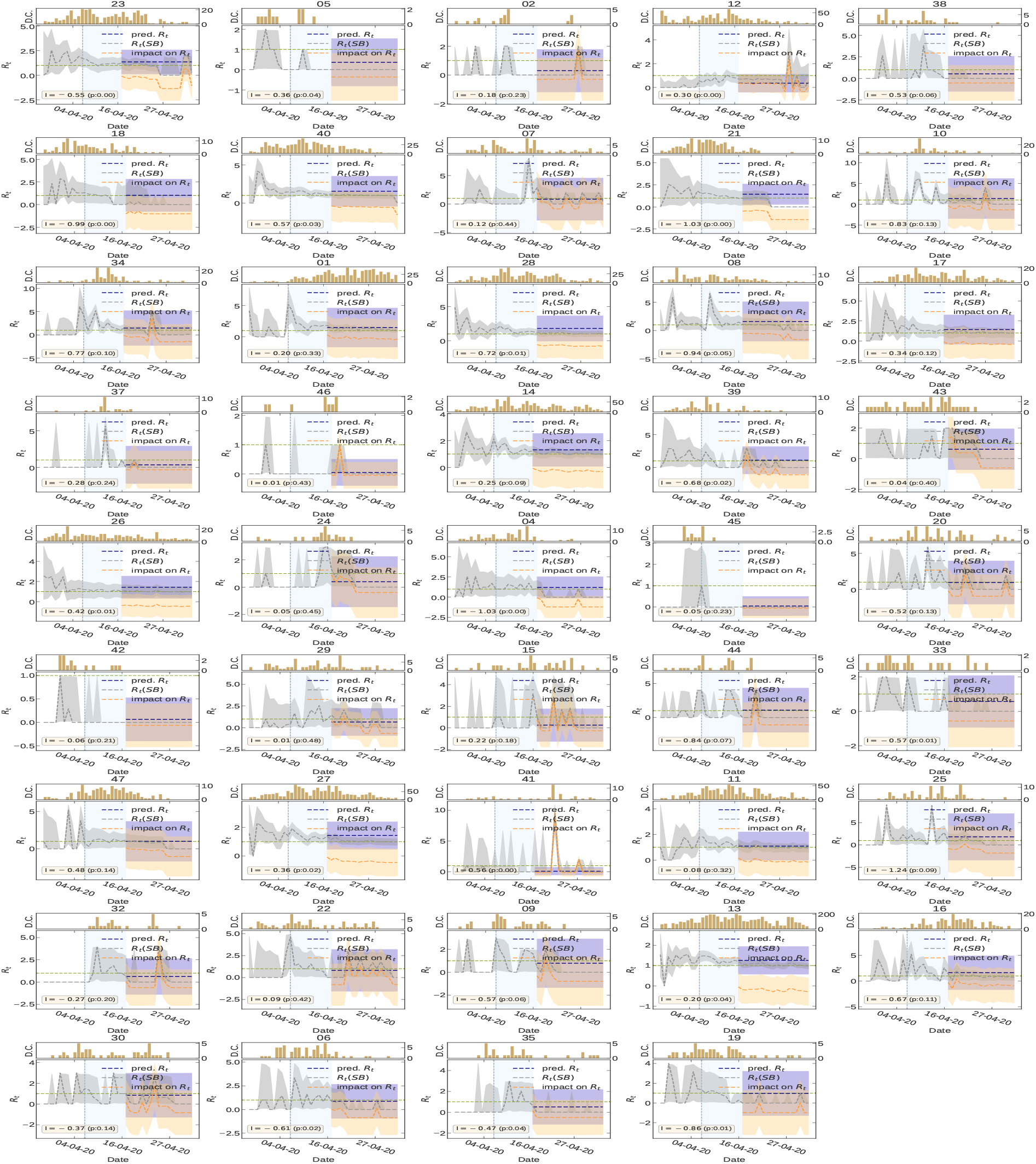
Impact of Governmental interventions on *R_t_* in different administrative divisions of Japan.

**Figure S18:**
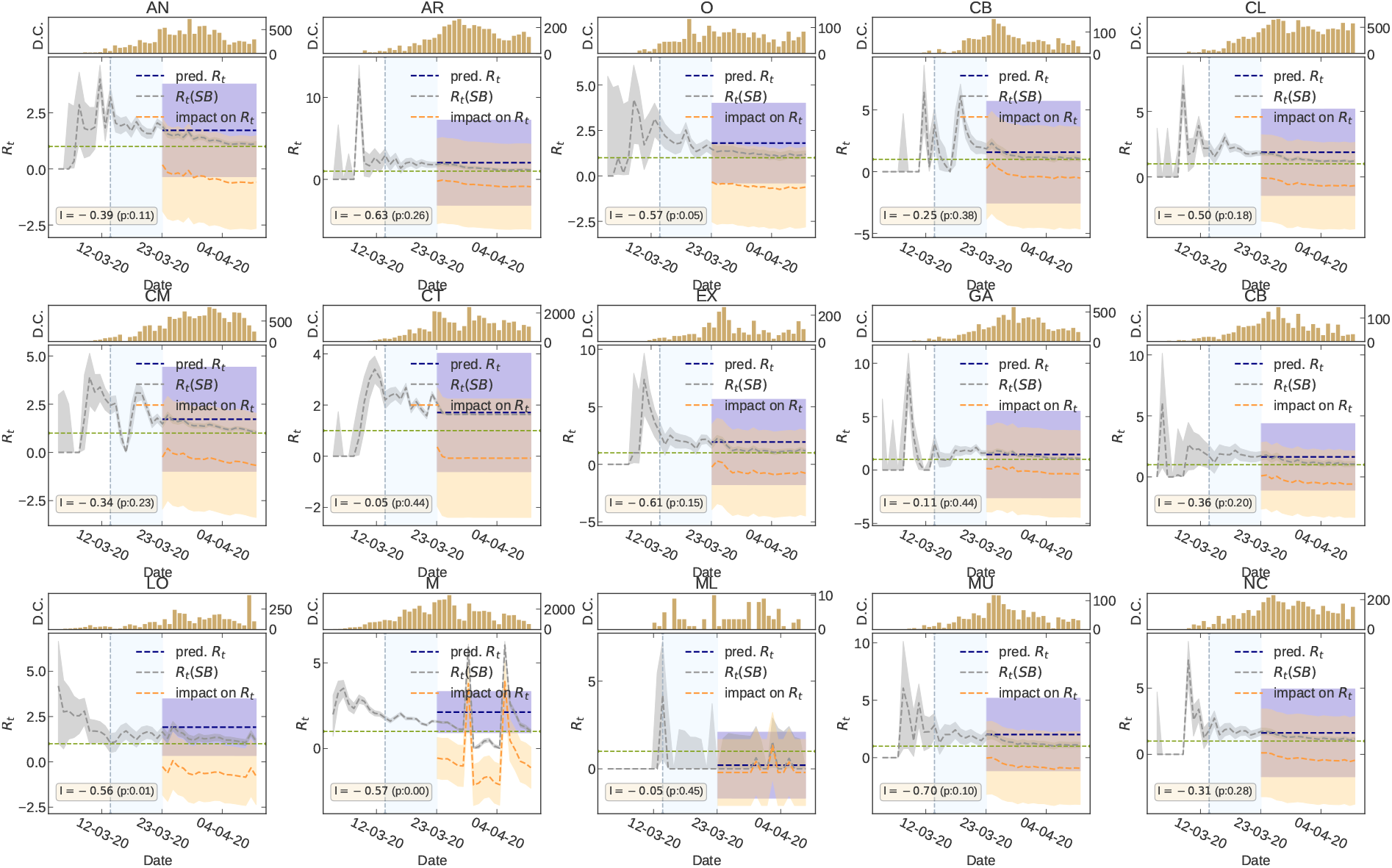
Impact of Governmental interventions on *R_t_* in different administrative divisions of Spain.

**Figure S19:**
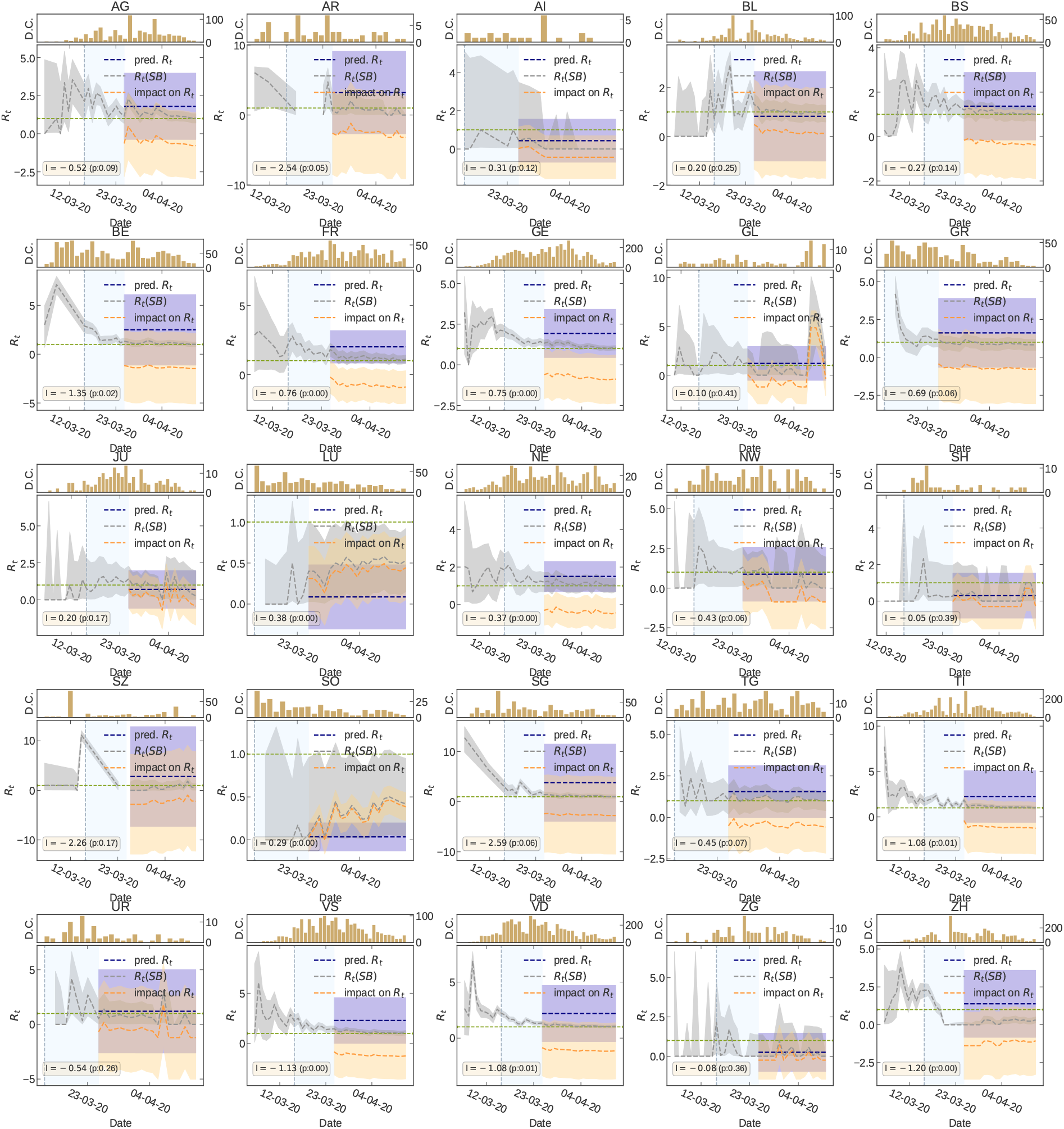
Impact of Governmental interventions on *R_t_* in different administrative divisions of Switzerland.

**Figure S20:**
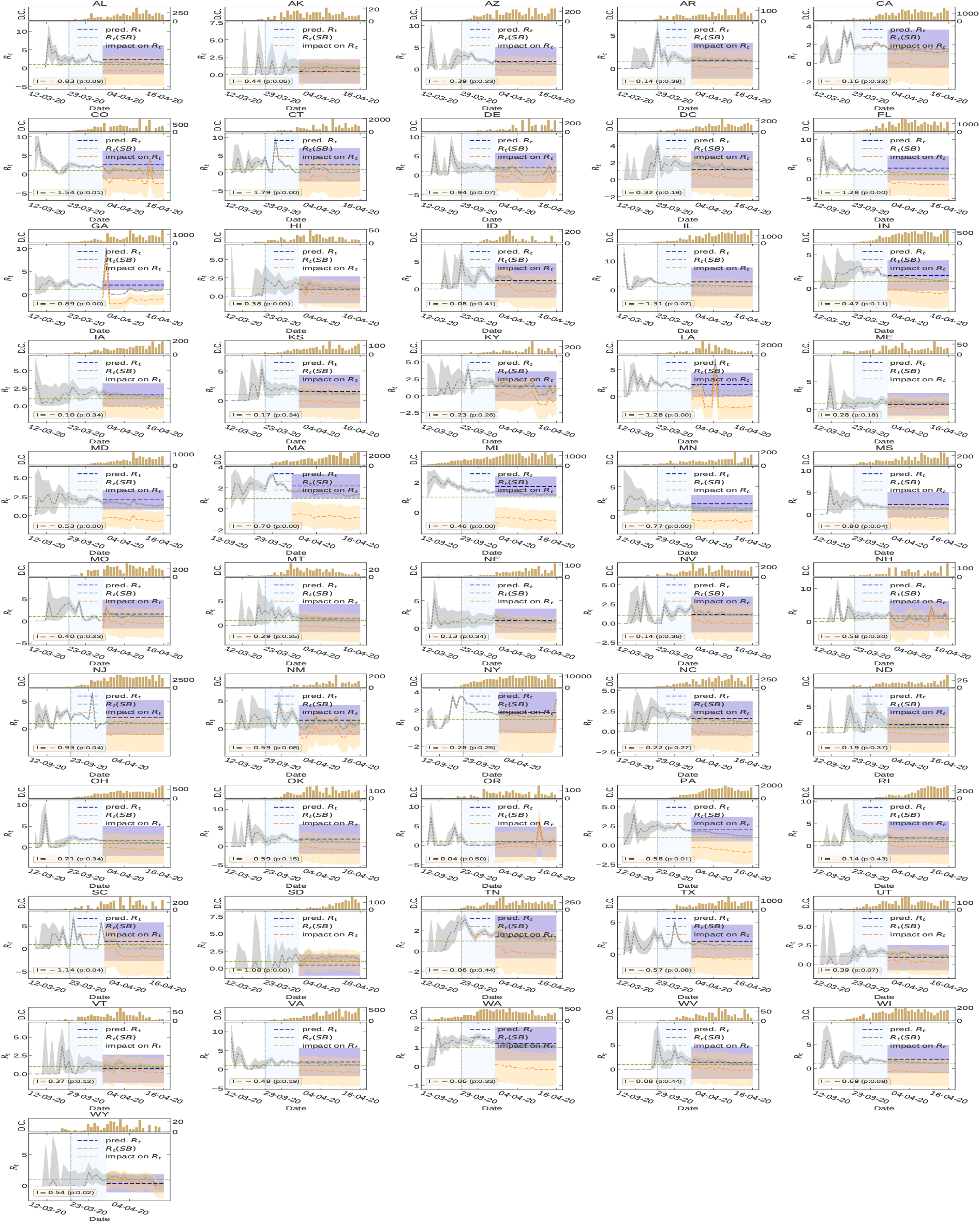
Impact of Governmental interventions on *R_t_* in different administrative divisions of United States.

## Notes

### Competing Interest Statement

The authors have declared no competing interest.

### Funding Statement

Not applicable.

